# Accelerometer-Derived and Genetically Inferred Physical Activity and Human Disease

**DOI:** 10.1101/2021.08.05.21261586

**Authors:** Shaan Khurshid, Lu-Chen Weng, Victor Nauffal, James P. Pirruccello, Rachael A. Venn, Mostafa A. Al-Alusi, Emelia J. Benjamin, Patrick T. Ellinor, Steven A. Lubitz

## Abstract

Physical activity is favorable to health but the relations with human disease and causal effects are poorly quantified. Previous studies have largely relied on self-reported estimates^1–3^ which are subject to recall bias, confounding, and reverse causality. Using wrist-worn accelerometer measurements from the UK Biobank, we tested associations between moderate-to-vigorous physical activity (MVPA) – both total MVPA minutes and whether MVPA was above a guideline-based threshold of ≥150 minutes/week^4–6^ – and incidence of over 1,200 diseases. In 96,466 adults who wore accelerometers for one week (mean age 62±8 years), MVPA was associated with 401 (33%) tested diseases at a false discovery rate of 1% during a median of 6.2 years follow-up. Greater MVPA was overwhelmingly associated with reduced disease risk (98% of associations) with hazard ratios (HRs) ranging from 0.10-0.95 per standard deviation (SD) and associations spanning all 18 disease categories tested. A similar pattern of associations was observed when assessing the guideline-based threshold of ≥150 MVPA minutes/week. We examined a polygenic risk score for MVPA as an instrumental variable for activity within a separate UK Biobank sample (N=392,058, mean age 57±8 years). Greater genetically inferred MVPA was associated with reduced risk of 182 (14%) incident diseases (HR range 0.74-0.97 per 1 SD), and included strong associations conferring reduced risks of obstructive bronchitis, peripheral vascular disease, type 2 diabetes, and gastritis/duodenitis. Objective physical activity is broadly associated with lower disease incidence and many associations are consistent with a causal effect.

---

Physical activity may have important health benefits,^5^ though the extent and causal effects of physical activity on human disease are poorly quantified. Examining relations between activity and disease risk may provide a comprehensive understanding of the benefits of physical activity, a modifiable lifestyle behavior. Past studies have generally been small and limited in power, and assessed physical activity using self-report data,^1–3^ which is subject to recall bias and correlates only modestly with measured energy expenditure.^7^ Studies that have measured activity objectively have frequently employed proprietary step counts^8^ or continuous acceleration,^9^ which may be difficult to contextualize in terms of consensus activity recommendations, which typically recommend specific quantities of moderate-to-vigorous physical activity (MVPA).^4–6^ Regardless of the measurement method, traditional study designs may remain subject to confounding (e.g., frailty)^10^ and reverse causality.

To address these challenges, we examined a unique, large prospective cohort study, the UK Biobank, which comprises over 90,000 individuals who wore wrist-worn triaxial accelerometers for one week and who also underwent genome-wide genotyping. The use of wearable accelerometer-based physical activity measurements allowed for precise, objective, and reproducible^9^ ascertainment of physical activity, quantified in terms of MVPA and also classified into binary categories divided at a guideline-recommended threshold of ≥150 minutes/week.^4–6^ Since physical activity has a heritable component,^9^ we then conducted Mendelian randomization (MR) analyses using a genetic risk score as an instrumental variable to further assess potential causal relations between objective physical activity and disease risk in nearly 400,000 separate UK Biobank participants who did not contribute accelerometry data.^11^

## Results

### Measured activity sample

After removing individuals whose accelerometer measurements failed quality control metrics, we performed phenotypic association testing in 96,466 individuals (**Figure 1**). The mean age was 62±8 years and 56% were female. Individuals had a median MVPA of 135 minutes/week (quartile-1: 65, quartile-3: 250) and 46% of individuals achieved guideline-recommended levels. MVPA distributions are shown in **Supplemental Figure 1**. Detailed sample characteristics are shown in **Table 1** and an overview of the samples is depicted in **Supplemental Figure 2**.

**Table 1.** Baseline characteristics of study samples

| **Baseline Characteristic\*** | **Wrist-worn accelerometer sample (N=96,466)** | **Self-reported activity sample (N=457,633)<sup>†</sup>** | **Genetically inferred activity sample (N=392,058)** |
| --- | --- | --- | --- |
| Age | 62.4 ± 7.8 | 56.9 ± 8.1 | 57.1 ± 8.1 |
| Female sex | 54,317 (56.3%) | 245,681 (53.7%) | 210,793 (53.8%) |
| Race<sup>‡</sup> |  |  |  |
| White | 93,177 (96.6%) | 434,397 (94.9%) | 367,216 (93.7%) |
| Asian | 1,118 (1.2%) | 8,871 (1.9%) | 9,823 (2.5%) |
| Black | 807 (0.8%) | 6,559 (1.4%) | 6,861 (1.7%) |
| Mixed | 528 (0.5%) | 2,654 (0.6%) | 2,328 (0.6%) |
| Other/Unknown | 836 (0.9%) | 5,152 (1.1%) | 5,830 (1.5%) |
| Tobacco |  |  |  |
| Current | 6,496 (6.7%) | 46,191 (10.1%) | 44,345 (11.3%) |
| Former | 34,721 (36.0%) | 159,252 (34.8%) | 134,119 (34.2%) |
| Never | 55,249 (57.3%) | 252,190 (55.1%) | 213,594 (54.7%) |
| Height (cm) | 169.0 ± 9.1 | 168.7 ± 9.3 | 168.3 ± 9.3 |
| Weight (kg) | 76.6 ± 15.4 | 78.0 ± 15.9 | 78.4 ± 16.0 |
| Body mass index (kg/m<sup>2</sup>) | 26.7 ± 4.5 | 27.3 ± 4.7 | 27.6 ± 4.8 |
| Hypertension | 27,154 (28.1%) | 130,776 (28.6%) | 118,170 (30.1%) |
| Diabetes | 3,205 (3.3%) | 11,594 (2.5%) | 11,426 (2.9%) |
| *Baseline characteristics defined at end accelerometer wear for acceleration sample, otherwise defined at enrollment<br><sup>†</sup>N=354,083 are included in the genetically inferred activity sample and N=92,289 are included in the wrist-worn accelerometer sample. There is no overlap between the accelerometer sample and the genetic inferred activity sample (see **Supplemental Figure 2**).<br><sup>‡</sup>Self-reported race |  |  |  |

**Figure 1.**
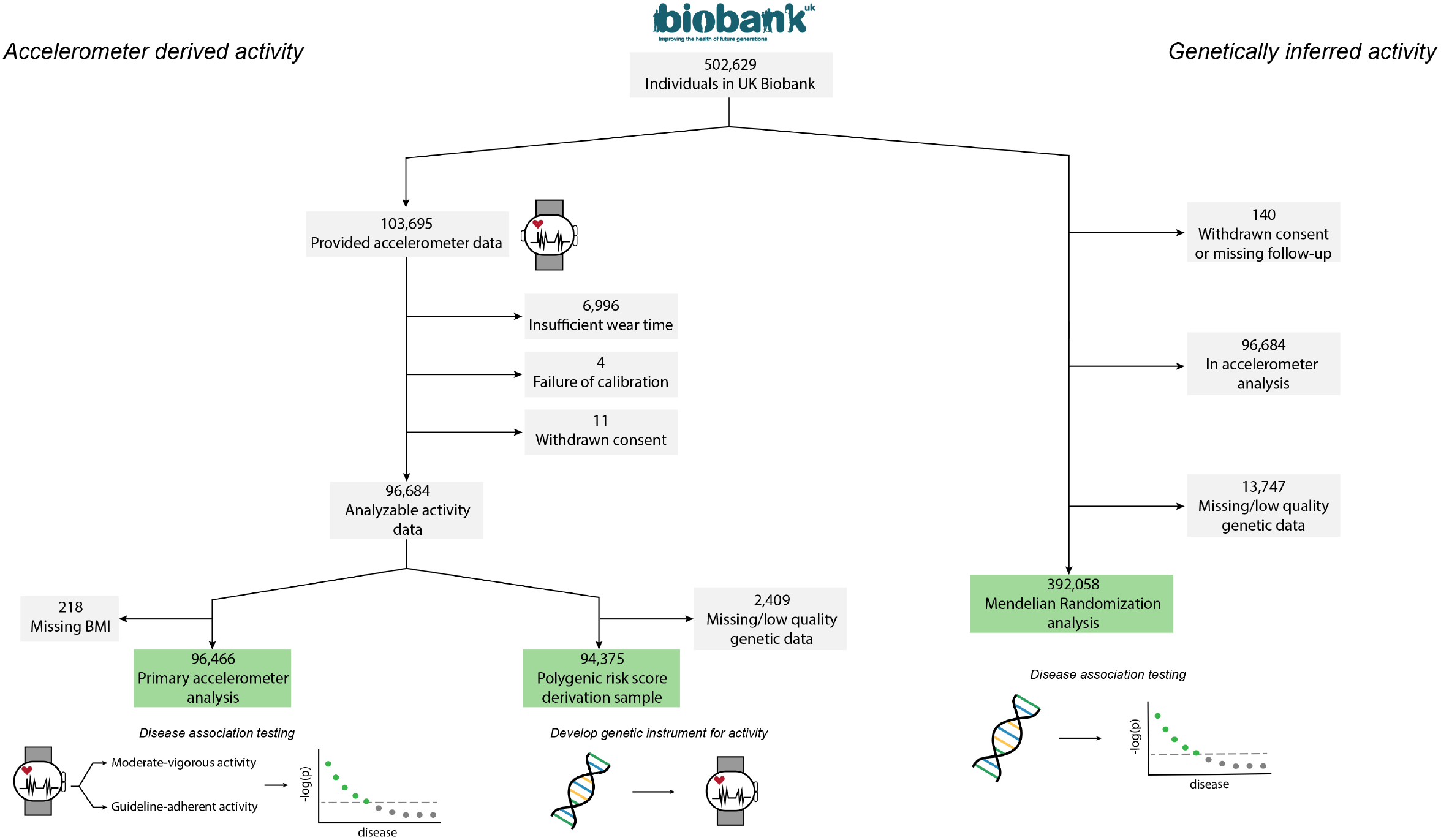
Study overview Depicted is a graphical overview of the study design. We performed association testing in 96,466 UK Biobank participants who wore a wrist-worn accelerometer for one week. We then performed a genome-wide association study using this sample to develop a polygenic risk score to act as a genetic instrument for activity. In a separate set of 392,058 UK Biobank participants, we inferred physical activity using the genetic instrument and performed analogous association testing with incident disease.

### Associations between measured activity and incident disease

At a median follow-up of 6.2 years (5.7, 6.7), MVPA was associated with risk of 401 incident diseases at a false discovery rate (FDR) of 1% (**Figure 2a**). Of the significant associations, 394 (98%) indicated a decreased risk of disease with greater MVPA (hazard ratio [HR] range 0.10-0.95 per 1-standard deviation [SD]). Some of the strongest associations included substantially decreased risks of incident heart failure (HR 0.63, 95% CI 0.56-0.71), stage 3 chronic kidney disease (HR 0.66, 95% CI 0.60-0.73), sleep apnea (HR 0.66, 95% CI 0.59-0.75), and hyperlipidemia (HR 0.87, 95% CI 0.84-0.90) (**Supplementary Figure 3**). Overall, associations with reduced disease risk were enriched for cardiac (17%), digestive (14%), endocrine/metabolic (11%), and respiratory conditions (8%) (chi-square p<0.01). The distribution of effect sizes varied by disease category, with the lowest median hazard ratios observed for respiratory, endocrine/metabolic, and neurological diseases (**Figure 2b**). There were seven associations between greater MVPA and increased risk of disease (HR range 1.10-1.31), and all represented injuries/poisonings, musculoskeletal, and dermatologic conditions, including greater risk of disorders of muscle, ligament, and fascia (HR 1.10, 95% CI 1.03-1.17) and fracture of the radius or ulna (HR 1.11, 95% CI 1.04-1.18). We obtained similar results when categorizing MVPA at the ≥150 minutes per week threshold recommended in consensus guidelines^4–6^ (358 associations with decreased risk of disease, HR range 0.21-0.91, **Supplemental Figure 4**).

**Figure 2.**
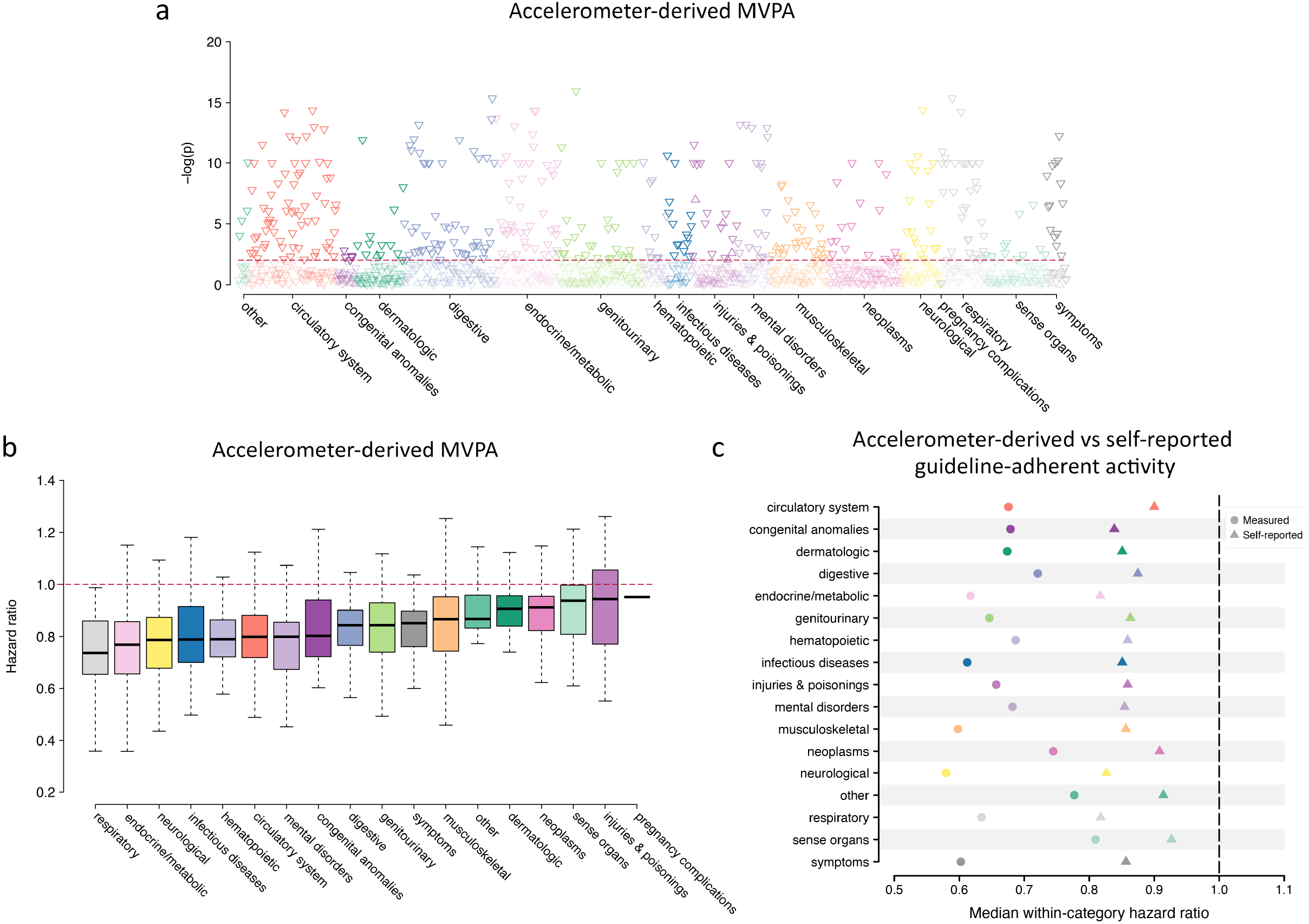
Measured physical activity and incident disease **Panel a** plots the negative log10 p-value for the association between accelerometer-derived moderate-to-vigorous physical activity (MVPA) and incident disease (grouped by category on the x-axis) in Cox proportional hazards models adjusted for age, sex, and body mass index, with darker shaded points meeting significance at a false discovery rate (FDR) of 1% (horizontal red line). Upward facing triangles represent increased risk (hazard ratios >1), while downward facing triangles represent reduced risk (hazard ratio <1). **Panel b** shows the distribution of hazard ratios per 1 standard deviation increase in MVPA observed across each disease category (x-axis), with the thick horizontal line depicting the within-category median hazard ratio, the box representing quartile 1 to quartile 3, and the whiskers extending 1.5 interquartile ranges beyond. Categories are arranged by increasing median hazard ratio, from lowest (left) to highest (right). **Panel c** compares the median within-category hazard ratio observed with guideline-adherent activity (i.e., ≥150 minutes of MVPA per week^4–6^) defined using accelerometer-derived (circles) versus self-reported (triangles) MVPA. Included in the comparison are 324 diseases for which there was a nominally significant association (p<0.05) with both exposure definitions and an effect size suggesting a decreased risk of disease.

### Secondary activity exposures

We also assessed disease associations using overall mean acceleration, which demonstrated a similar pattern of associations as accelerometer-derived MVPA (**Supplemental Figure 5**). Within 457,633 UK Biobank participants providing self-reported activity data (**Table 1** and **Supplemental Figure 2**), associations between self-reported MVPA (both continuous and categorized at the guideline-based threshold) were qualitatively similar, but effect sizes were consistently smaller (**Figure 2c** and **Supplemental Figures 6-7**). As compared to accelerometer-derived MVPA, self-reported activity had more associations indicating an increased risk of disease, primarily musculoskeletal conditions (e.g., osteoarthrosis, HR 1.07, 95% CI 1.06-1.08) and injuries/poisonings (e.g., joint/ligament sprain, HR 1.10, 95% CI 1.07-1.14) (**Supplemental Figures 8-9**). Full results of association testing using each of the outlined exposures are available in the **Supplemental Data**.

### Genome-wide association study of MVPA and development of genetic instrument

We then performed a genome-wide association study of square root-transformed MVPA in the UK Biobank accelerometer sample. We observed two loci meeting genome-wide significance, with the dominant locus previously reported for accelerometer-derived mean acceleration^12,13^ (**Supplemental Figures 10-11** and **Supplemental Table 1**). Total observed scale heritability (h2) was 0.11 (standard error 0.01). We then pruned the genetic association results to aggregate the association signals into a polygenic risk score (PRS), which served as a genetic instrument for physical activity. The PRS comprised 561 genetic variants and explained 6.6% of the variation in square root MVPA in a linear regression of square root MVPA on PRS, consistent with a sufficiently strong instrument (F-score: 6,716; p<0.01, **Supplemental Table 2**).

### Genetically inferred activity sample

We then used the PRS as a genetic instrument to infer MVPA levels in a separate sample of 392,058 UK Biobank participants who did not contribute to the accelerometer sub-study. The mean age was 57±8 years and 54% were female. Individuals had a median genetically inferred MVPA of 165 minutes/week (quartile-1: 128, quartile-2: 208) and 60% of individuals had genetically inferred MVPA achieving guideline-recommended levels. Inferred MVPA distributions are shown in **Supplemental Figure 1**. Detailed characteristics of the inferred activity sample are shown in **Table 1** and an overview of the sample is depicted in **Supplemental Figure 2**.

### Associations between genetically inferred activity and incident disease

At a median follow-up of 11.9 years (11.1, 12.6), genetically inferred MVPA was associated with 182 incident diseases (**Figure 3a**). All 182 associations demonstrated lower disease risk with greater inferred MVPA. The distribution of disease categories in which significant associations were observed was similar to that of measured MVPA, again with enrichment for digestive, circulatory, endocrine/metabolic, and respiratory conditions (**Figure 3a**). Some of the strongest associations included substantially decreased risks of pulmonary hypertension (HR 0.80, 95% CI 0.70-0.93), stage 3 chronic kidney disease (HR 0.94, 95% CI 0.91-0.97), sleep apnea (HR 0.93, 95% CI 0.90-0.97), and type 2 diabetes with neurological manifestations (HR 0.81, 95% CI 0.73-0.81) (**Supplementary Figure 3**).

**Figure 3.**
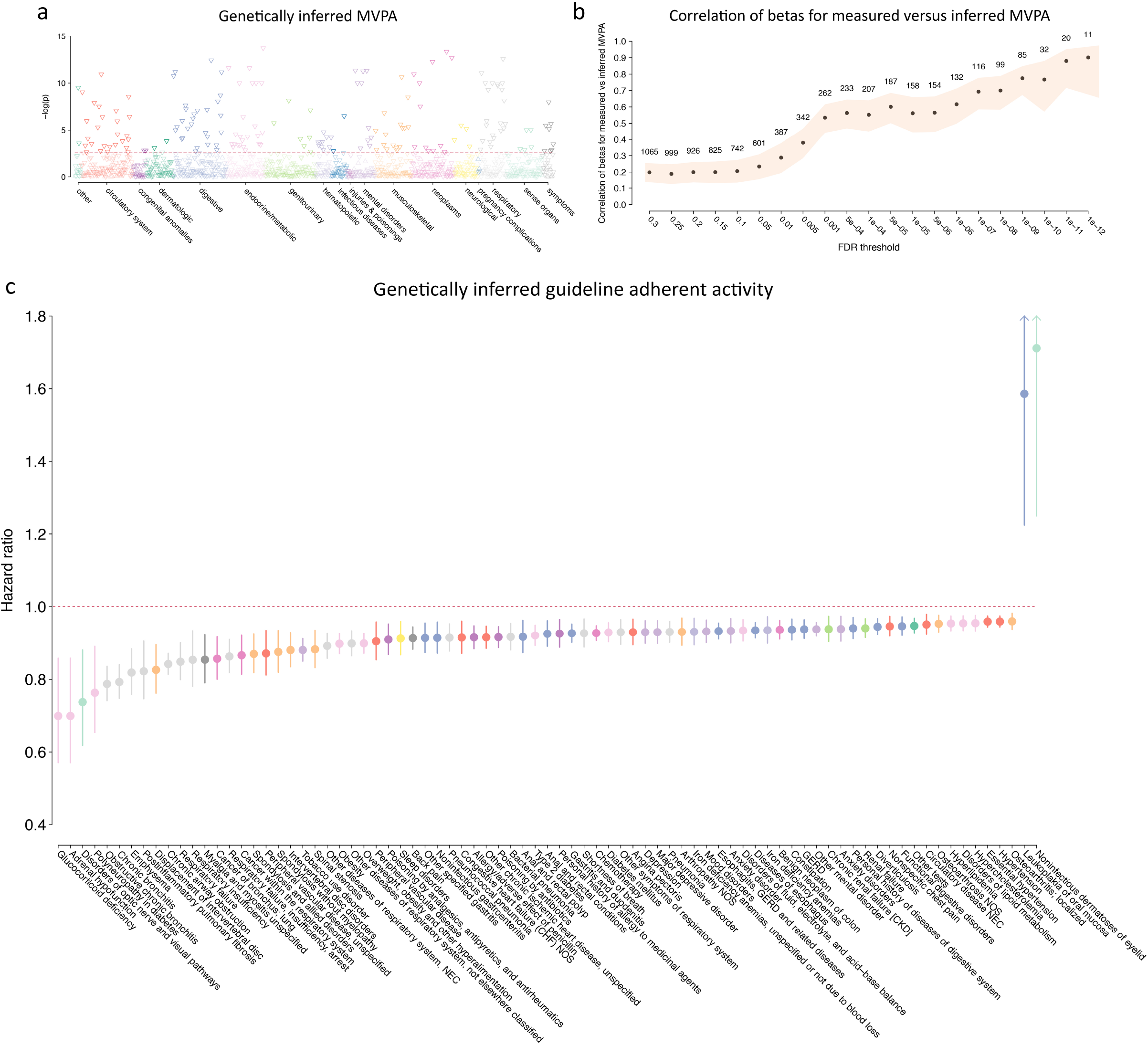
Genetically inferred activity and incident disease **Panel a** plots the negative log10 p-value for the association between genetically inferred MVPA and individual diseases (grouped by category on the x-axis), in Cox proportional hazards models adjusted for age and sex, with darker shaded colors representing significant associations at a false discovery rate (FDR) of 1% (threshold depicted by horizontal red line). **Panel b** shows the Pearson correlation between disease-specific beta coefficients for accelerometer-derived versus genetically inferred MVPA from their respective Cox models, as a function of increasing false discovery rate (FDR) threshold used to define a significant association with accelerometer-derived MVPA (x-axis). The number of diseases included in the correlation assessment is indicated above each point, and the 95% confidence interval is depicted by the shaded area. **Panel c** shows the hazard ratios associated with genetically inferred guideline-adherent activity (i.e., ≥150 minutes of inferred MVPA per week^4–6^), with specific diseases listed on the x-axis. Points are colored to match the disease category groupings used in **Panel a**. Confidence intervals ending with arrows extend beyond the plotting area but have been truncated for graphical purposes.

Guideline-adherent activity according to genetically inferred MVPA was associated with reduced risk of 79 diseases (98% of all significant associations, **Figure 3d** and **Supplemental Figure 12**). Disease-specific effect sizes between genetically inferred and accelerometer-derived MVPA were substantially correlated, with increasing strength of correlation among diseases more strongly associated with measured MVPA (maximum correlation r=0.90, 95% CI 0.66-0.97, **Figure 3c**).

In total, 70 diseases were significantly associated with both measured (in the accelerometer sample) and inferred (in the non-accelerometer sample) guideline-adherent activity (**Table 2**). Disease-specific effect sizes were highly correlated (r=0.83, 95% CI 0.75-0.89). Significant genetic associations with some of the largest effect sizes included obstructive chronic bronchitis (HR 0.79, 95% CI 0.74-0.84, p=1.6×10^−15^), peripheral vascular disease (HR 0.90, 95% CI 0.85-0.96, p=6.0×10^−12^), type 2 diabetes (HR 0.92, 95% CI 0.89-0.95, p=1.0×10^−10^), and gastritis/duodenitis (HR 0.93, 95% CI 0.90-0.95, p=3.8×10^−5^). Plots of the 5-year cumulative risk of these four conditions demonstrated consistent and substantial separation of longitudinal disease incidence on the basis of both measured and genetically inferred guideline-adherent activity (**Figure 4**). Risk curves adjusted for age and stratified by sex showed a similar pattern (**Supplemental Figures 13-14**). Full results of association testing using the genetically inferred MVPA exposures are available in the **Supplemental Data**.

**Table 2.**
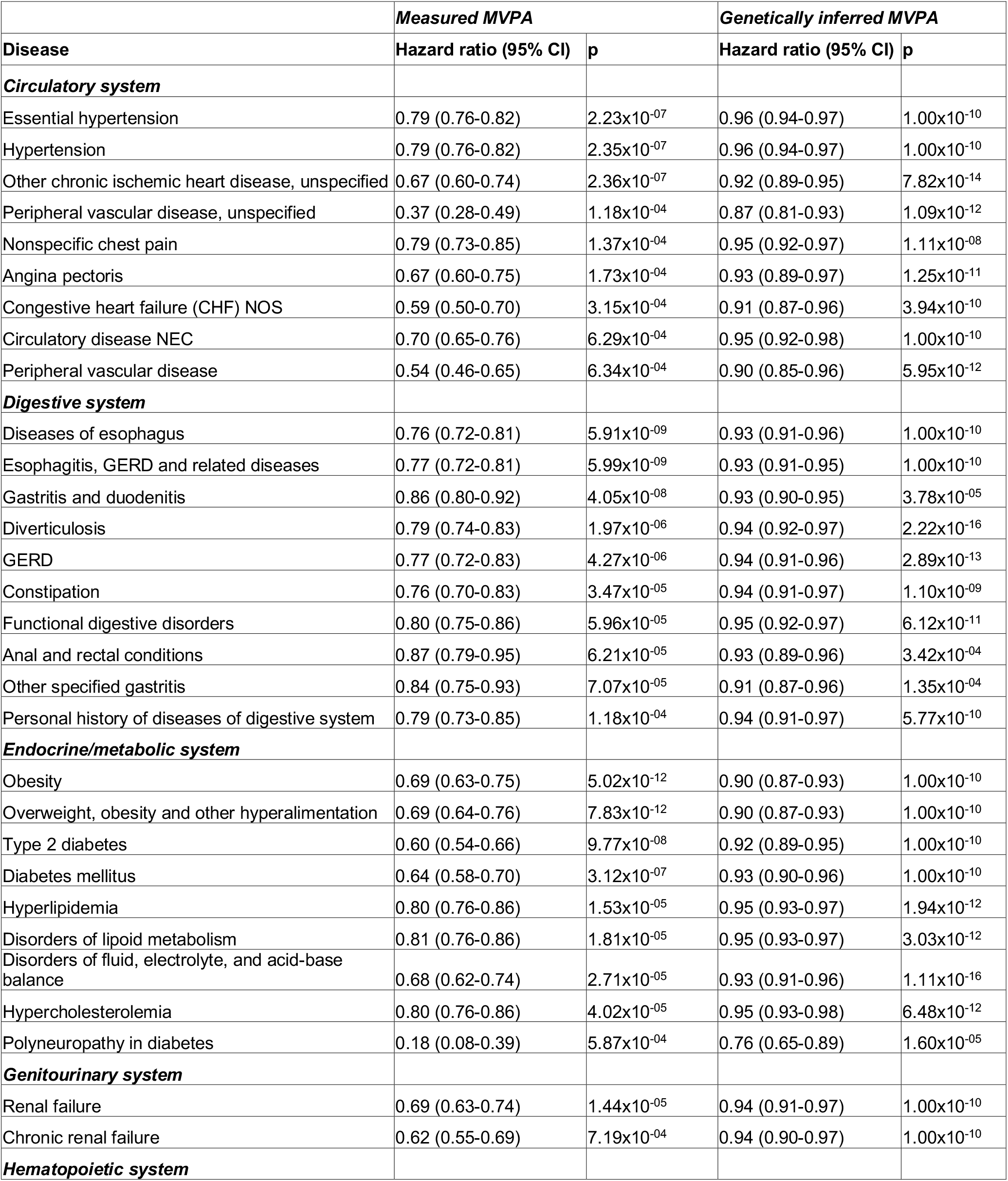
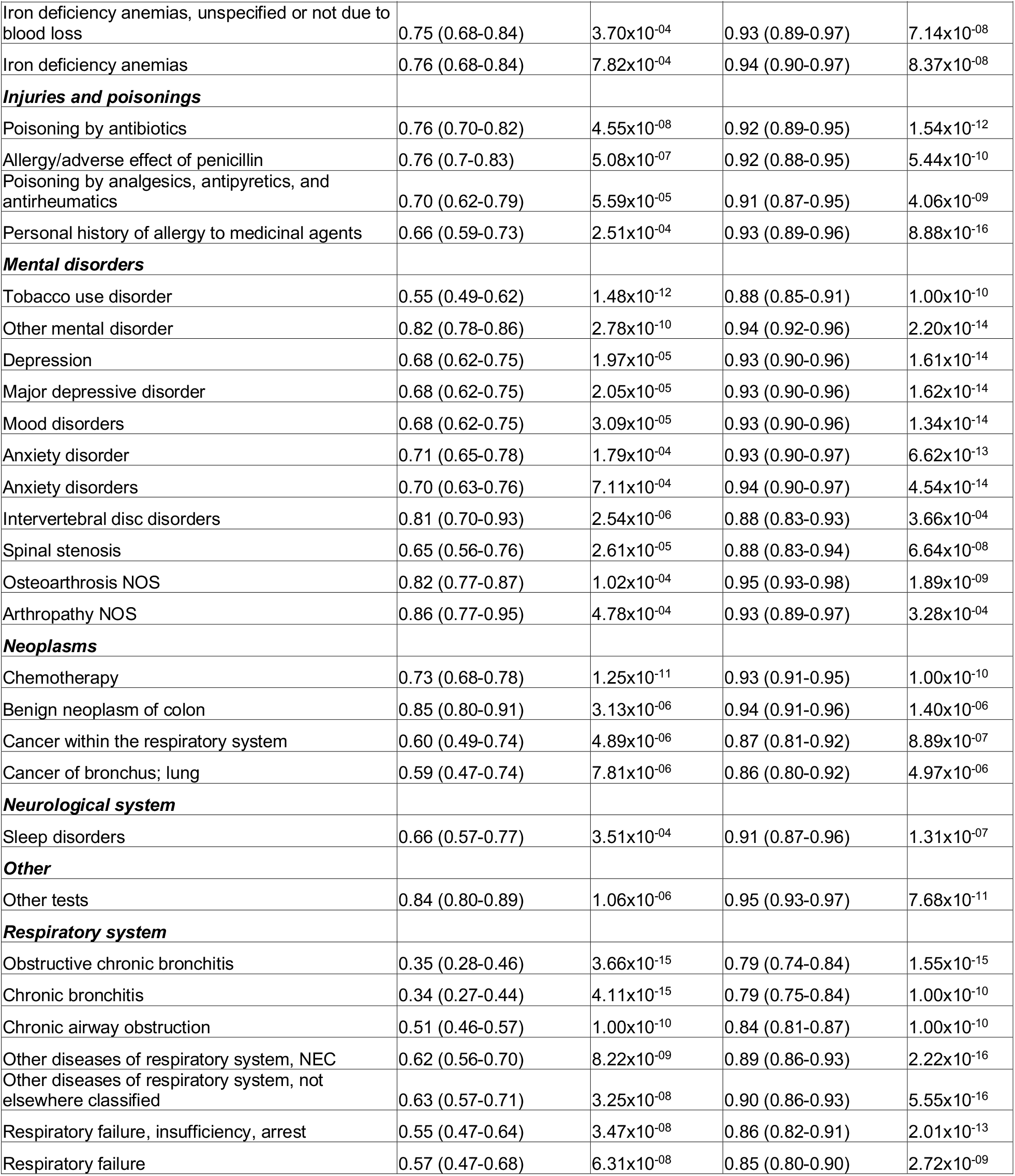
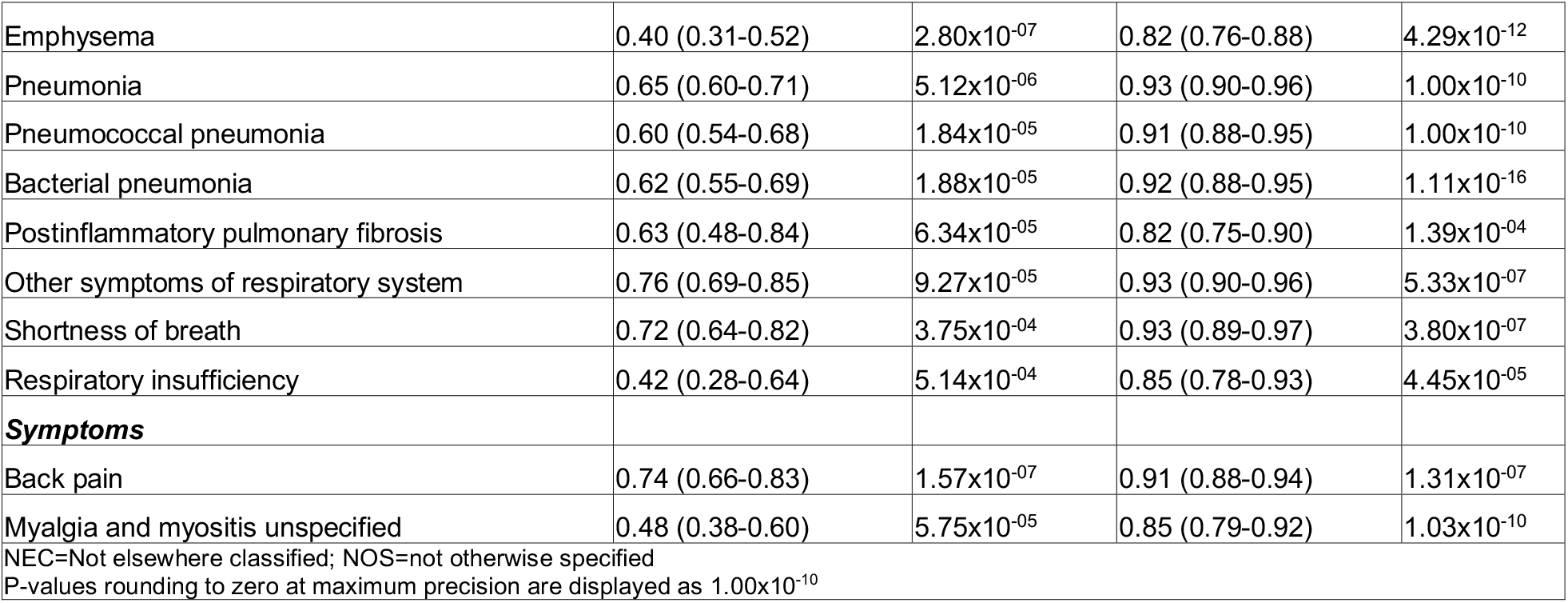
Diseases associated with at least 150 minutes per week of moderate to vigorous physical activity

**Figure 4.**
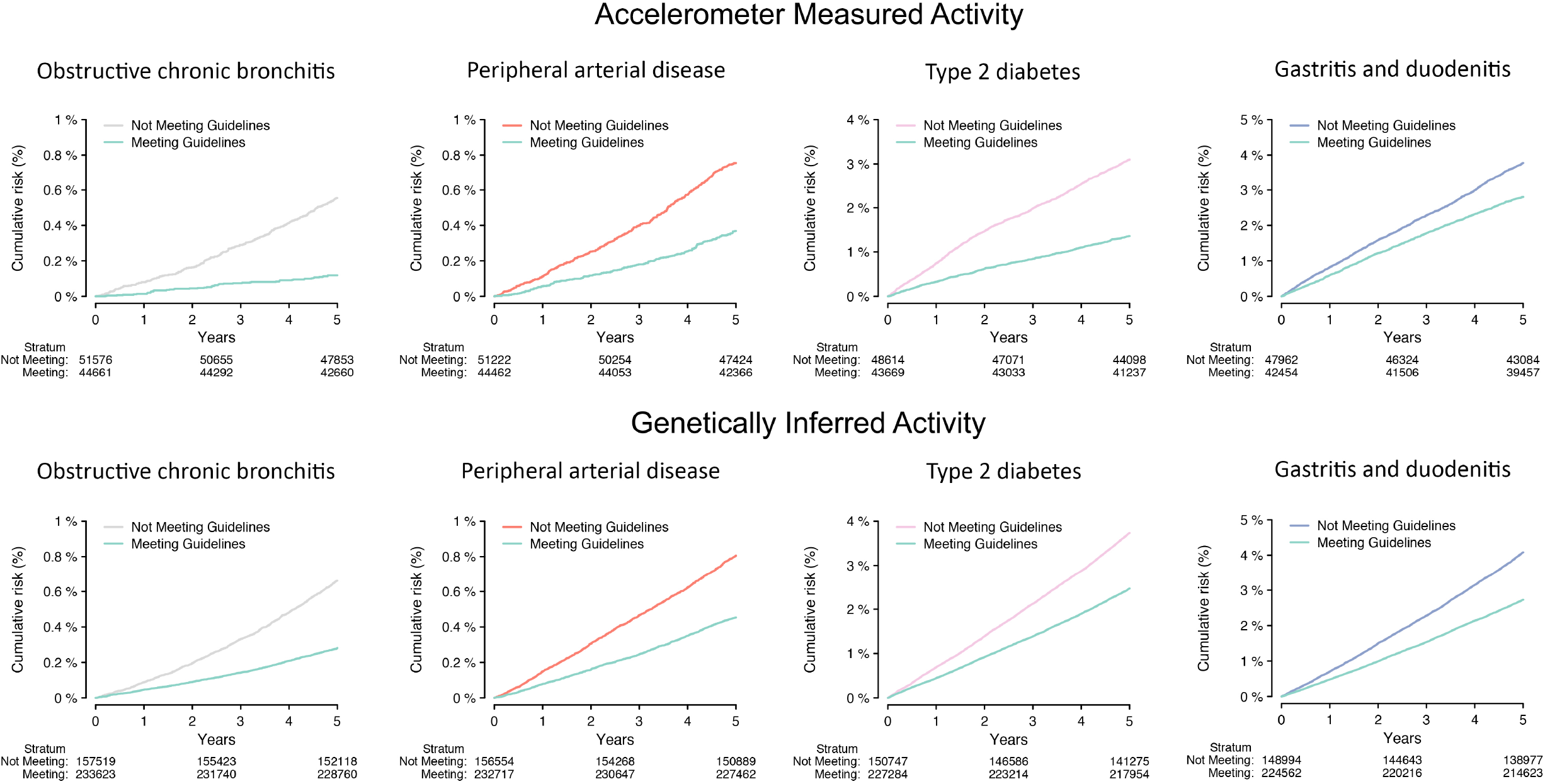
Cumulative risk of disease stratified by guideline-adherent physical activity Depicted is the 5-year cumulative risk of obstructive chronic bronchitis, peripheral arterial disease, type 2 diabetes, and gastritis/duodenitis, stratified guideline-adherent activity according to accelerometer-derived MVPA (top panels) and according to genetically inferred MVPA (bottom). In each plot, individuals are grouped into binary categories according to the guideline-based threshold of ≥150 minutes of MVPA/week,^4–6^ and the number remaining at risk over time is depicted below. Representative diseases were selected from the four categories having the greatest enrichment for associations with activity, where each disease was significantly associated with both accelerometer-derived and genetically inferred activity at a false discovery rate of 1%.

### Secondary analyses

Associations between greater measured and genetically inferred MVPA and decreased risk of disease were consistently observed across subgroups of age (i.e., age <55, age 55-64, and age ≥65), although both the number of significant associations and strength of effect sizes were greatest in older individuals (**Supplemental Figures 15-16**). When assessing alternative MVPA thresholds, a similar pattern of associations was observed at both ≥75 and ≥300 MVPA minutes/week, but the total number of significant associations at 300 MVPA minutes/week was smaller (**Supplemental Figures 17-18**). Associations between greater genetically inferred MVPA and lower risk of chronic obstructive bronchitis, peripheral vascular disease, type 2 diabetes, and gastritis/duodenitis were each similar when assessed using a genetic instrument derived after excluding individuals with any of the four conditions prevalent at the time of blood sample collection, suggesting that such associations were not driven by reverse causality (**Supplemental Table 3**).

## Discussion

In summary, within over 90,000 individuals wearing a wrist-based activity sensor over the course of one week, we quantified associations between objective physical activity (quantified as MVPA) and future risk of over 1,000 diseases. We observed strong associations between physical activity and hundreds of conditions, in which activity was linked to reduced risks of cardiac, digestive, endocrine/metabolic, and respiratory conditions among other disease. Using a genetic instrument for objective physical activity within nearly 400,000 separate individuals, we observed associations between physical activity and over 180 disease entities which are overwhelmingly consistent with a causal protective mechanism.

Our findings support previous observations that objective activity levels are a powerful indicator of disease risk, and are potentially more informative than self-reported activity. Multiple studies have suggested that self-reported activity may be biased,^14^ and therefore a weak surrogate for measured activity.^7^ Indeed, recent evidence suggests that objective activity measures may be a more powerful predictor of cardiovascular outcomes^15^ and mortality^14^ than self-reported activity within the same population. We similarly observed substantially stronger effect sizes when assessing guideline-adherent activity using accelerometer-derived as opposed to self-reported MVPA. Interestingly, although the distribution of significant associations was qualitatively similar using objective versus self-reported data, self-reported data suggested a considerably greater number of associations indicating increased disease risk, primarily comprising musculoskeletal conditions and injuries/poisonings. Future work is warranted to better understand potential differences in information content between objectively measured and self-reported physical activity, and how such differences may impact efforts to modify disease risk.

Our results suggest that interventions to increase adherence to guideline-recommended activity levels assessed by objective measurement will have a wide-ranging favorable effect on future disease risk. Whether assessed as a continuous variable or dichotomized at a guideline-recommended threshold, greater MVPA was associated with reduced risk of roughly 400 diseases. Although associations were enriched for cardiac, digestive, endocrine/metabolic, and respiratory conditions, the breadth of associations we observed spanned every disease category tested, including several protective associations involving neurologic, respiratory, and psychiatric conditions. For example, we observed that individuals meeting guideline-recommended levels of accelerometer-derived MVPA had nearly 40% lower risk of an incident sleep disorder, which is consistent with randomized trial evidence supporting a beneficial role of exercise on sleep.^16,17^ In secondary analyses we also observed that achievement of lower levels of activity (i.e., ≥75 minutes of MVPA/week) was also associated with lower disease risk, suggesting that efforts to increase objective activity – even if not to the point of achieving guideline-recommended levels – may be broadly beneficial. Although our findings suggest that greater activity should prevent disease, we observed a small number of associations between physical activity and increased risk of certain conditions. In particular, physical activity may increase the likelihood of developing certain musculoskeletal disorders, injuries, and dermatologic diseases (e.g., corns and calluses), potentially reflecting use-related degeneration.^18^ On balance, however, our findings suggest that future efforts to optimize objective activity have potential for substantial and wide-ranging beneficial effects spanning the full spectrum of human disease.

The results of the MR analyses, a form of instrumental variable analysis, indicates that increased activity may be causally associated with reduced disease risk. For certain conditions (e.g., obstructive chronic bronchitis, type 2 diabetes), individuals meeting guideline-recommended activity levels (categorized using either accelerometer-derived or genetically-inferred MVPA) had a substantially lower risk of disease over several years. Some of the associations we observed confirm causal patterns that are relatively well-understood, such as greater activity leading to decreased diabetes risk through mechanisms such as reduced adiposity^19^ and increased insulin sensitivity.^20^ Similarly, previous work supports a causal effect of exercise on decreased burden of atherosclerosis, potentially mediated by decreased vascular inflammation^21^ and improved endothelial function.^22^

Yet we also observed associations between genetically inferred MVPA and conditions for which causal pathways are not well-established, including certain pulmonary (e.g., obstructive chronic bronchitis, chronic airway obstruction, emphysema, and pneumonia) and digestive (e.g., gastritis/duodenitis, diverticulosis) conditions. Although physical activity has been identified as a potential risk factor for recurrent exacerbations of pulmonary disease,^23^ and exercise may modulate risk of gastritis through beneficial effects on stress, acid secretion, and chronic inflammation,^24,25^ future work is needed to better clarify potential mechanisms. Nevertheless, our findings provide important causal evidence that physical activity may lower disease risk.

Our study should be considered in the context of design. First, objective physical activity was measured for only one week. It is possible that a longer duration of monitoring would have led to more accurate classification of activity. Second, follow-up time after accelerometry (approximately six years) remains fairly limited. Third, given our intent to broadly survey the entire disease phenome for potential causal associations between physical activity and disease, we utilized a PRS as a genetic instrument in a one-sample Mendelian randomization design, which may be prone to bias introduced by horizontal pleiotropy that we could not account for. Reassuringly, several of the associations we identified are consistent with previously established causal mechanisms.^16,17,19–22,26^ Furthermore, associations with exemplar diseases remained robust in sensitivity analyses utilizing a genetic instrument developed after excluding individuals with those diseases present at the time of blood sample collection, providing evidence against reverse causation affecting the genetic instrument. Fourth, the PRS we utilized was derived from and implemented in a source population primarily of European ancestry. As a result, the strength of the genetic instrument may vary if deployed in different populations. Fifth, although our genetic instrument was comparably strong,^27^ lack of associations with specific diseases may be related to insufficient power. Indeed, we observed that disease-specific effect sizes were correlated, even for diseases not significantly associated with inferred MVPA at our relatively stringent FDR threshold of 1%. Sixth, our results were obtained within a single community-based sample of primarily European ancestry and may not generalize to other populations.

In summary, we performed phenome-wide association testing for incident disease using a unique resource of objectively measured physical activity obtained within over 90,000 individuals. We observed that objective activity – defined as continuous MVPA as well as according to guideline-based thresholds – is associated with reduced risk of roughly 400 incident conditions spanning the full spectrum of human disease. Using a genetic instrument for objective activity, we found evidence of causal associations between greater activity and reduced risk of over 180 diseases enriched for cardiac, digestive, endocrine/metabolic, and respiratory conditions. Our findings prioritize optimization of objective physical activity levels as an important population-level mechanism to broadly reduce risk of future disease.

## Methods

### Data availability

UK Biobank data are freely available for research purposes by application (https://www.ukbiobank.ac.uk/). Data processing scripts used to perform the analyses described herein can be found at (https://github.com/shaankhurshid/acceleration_phewas). Detailed summary data supporting major study results are provided in the **Supplementary Data**.

### Study population

The UK Biobank is a prospective cohort of 502,629 participants enrolled between 2006-2010.^28^ Briefly, 9.2 million individuals aged 40-69 years living within 25 miles of 22 assessment centers in the UK were invited, and 5.4% participated in the baseline assessment. Questionnaires and physical measures were collected at recruitment, and all participants are followed for outcomes through linkage to national health-related datasets. All participants provided written informed consent. The UK Biobank was approved by the UK Biobank Research Ethics Committee (reference# 11/NW/0382). Use of UK Biobank data (application 17488) was approved by the local Mass General Brigham Health Institutional Review Board.

### Accelerometer-derived physical activity

Between February 2013-December 2015, 236,519 UK Biobank participants were invited to wear a wrist-worn accelerometer for one week, of whom 106,053 agreed to participate and 103,695 submitted data.^9^Participants were sent an Axivity AX3 (Newcastle upon Tyne, UK) wrist-worn triaxial accelerometer. The sensor captured continuous acceleration at 100Hz with dynamic range of±8*g*.

As described previously, acceleration signals were calibrated to local gravity.^9,29^ Sample data were combined into 5-second epochs, with each epoch represented by the average vector magnitude. Non-wear time was identified as consecutive stationary episodes ≥60 minutes in duration in which all three axes had standard deviation <13.0m*g*.^30,31^ Epochs representing non-wear time were imputed based on the average of similar time-of-day vector magnitude and intensity distribution data points on different days. We excluded individuals with insufficient wear-time to support imputation (<72 hours of wear-time or no wear data in each one-hour period of the 24-hour cycle), and whose signals were insufficient for calibration.^9^

The primary accelerometer-derived exposure was minutes of moderate-to-vigorous physical activity (MVPA), defined as the sum of 5-second epochs where mean acceleration was ≥100m*g*.^32,33^ To reduce the likelihood of misclassifying artifact as MVPA, we extracted MVPA in bouts (5-minute periods where ≥80% of epochs met the MVPA threshold).^33,34^ We then classified whether MVPA levels met recommended levels cited in guidelines from the World Health Organization,^4^ American College of Cardiology/American Heart Association,^5^ and European Society of Cardiology.^6^ Secondary exposures included overall mean acceleration, a surrogate for global physical activity that has been validated against energy expenditure.^31,35^ We extrapolated the observed MVPA rate to one week for the 5,043 individuals (5%) contributing less than a full week of wear time to account for variable wear time and to facilitate categorization of weekly guideline-based activity.

### Self-reported physical activity

Self-reported physical activity data were obtained at enrollment using the short-form international physical activity questionnaire (IPAQ).^36^ We quantified self-reported activity as weekly minutes of MVPA to mirror our accelerometer-based analyses, which quantified total time spent performing activities of moderate or greater intensity.

### Polygenic risk score for physical activity

We then analyzed individuals in the accelerometer sample with genome-wide genotyping data to develop a polygenic risk score (PRS) for objective physical activity, which served as an instrumental variable for physical activity. Details of genotyping and quality control in the UK Biobank have been described in detail previously.^37^ We performed a genome-wide association study (GWAS) of MVPA by fitting a linear mixed model with square root-transformed MVPA as the outcome of interest and age, sex, array platform, and the first five principal components of genetic ancestry as covariates. We utilized a square root-transform to attenuate high outliers while preserving sufficient information content to infer MVPA using the genetic instrument. Models were fit using BOLT-LMM.^38^ Inflation was assessed using quantile-quantile plots and calculating the linkage-disequilibrium score regression intercept.^39^ Lead single nucleotide variants (SNVs) were the most significantly associated variants with the outcome within a 500kb window. The genomic control factor was 1.15 with a linkage-disequilibrium score regression intercept of 1.008, consistent with polygenicity rather than inflation (**Supplemental Figure 7**). We used a pruning and thresholding approach to construct a PRS to estimate genetically determined MVPA, testing four sets of score parameters (p-value thresholds 1×10^−4^ and 1×10^−6^ permuted against R^2^ values of 0.3 and 0.5), each using a 250kb sliding window. The PRS most predictive of square root-transformed MVPA was constructed using a p-value threshold of 1×10^−4^ and an R^2^ of 0.3, and explained 6.6% of the variance in square root-transformed MVPA (F-score 6,716), which then served as the instrumental variable for accelerometer-derived MVPA (**Supplemental Table 2**).

We performed a one-sample Mendelian randomization analysis using the two-stage method to examine the causal association between physical activity and incident diseases. In the first stage we fit a linear model within the acceleration GWAS sample with square root-transformed MVPA as the outcome and age, sex, the first five principal components of genetic ancestry, and the best performing PRS as covariates. We then used the fitted model to predict square root-transformed MVPA in 392,061 UK Biobank participants who did not participate in the accelerometer sub-study, and subsequently squared the inferred value to estimate the untransformed MVPA. In the second stage, we examined the association between incident disease and inferred MVPA using Cox-proportional hazards models adjusting for age, sex and the first five principal components of ancestry. As with accelerometer-derived MVPA, we examined inferred MVPA as both a continuous variable and according to the guideline-based threshold of ≥150 minutes/week.^4–6^

### Outcomes

We defined diseases using v1.2 of the Phecode Map,^40^ a set of 1,867 disease definitions arranged into clinically meaningful groups and identified using standardized sets of International Classification of Disease, 9^th^ and 10^th^ revision codes. Diagnostic code sources included hospital data through linkage to national health-related datasets, as well as outpatient general practitioner visit data through linkage to electronic health records. In our incident disease analyses of accelerometer-derived variables, person-time started at the end of accelerometer measurement and person-time ended at an event, death, or last follow-up, whichever came first. In our incident disease analyses of both self-reported and inferred activity, person-time started at enrollment (i.e., date of genetic data collection) and was otherwise constructed similarly. The date of last follow-up varied according to availability of linked health data and was therefore defined as March 31, 2021 for participants enrolled in England and Scotland, and February 28, 2018 for participants enrolled in Wales.

### Statistical analysis

Associations between accelerometer-derived MVPA and incident disease were assessed using Cox proportional hazards regression, with adjustment for age, sex, and body mass index (BMI). Secondary models were constructed using a) adherence to standard physical activity guidelines (≥150 minutes of MVPA/week^4–6^) on the basis of accelerometer-derived MVPA, b) overall mean acceleration, c) self-reported MVPA, and d) adherence to standard physical activity guidelines (≥150 minutes of MVPA/week^4–6^) on the basis of self-reported data, as exposures of interest. For the 1-sample MR analysis, an analogous model was constructed using inferred MVPA as the exposure of interest, in models adjusted for age, sex, and the first five components of genetic ancestry. To prevent model instability, only diseases with ≥20 events (accelerometer-derived MVPA models) and ≥100 events (genetically inferred MVPA models) were tested. To assess effect size consistency between accelerometer-derived and inferred MVPA, we calculated the Pearson correlation between disease-specific beta coefficients corresponding to accelerometer-derived and inferred MVPA in the respective Cox proportional hazards models described above within their respective independent samples, as a function of the FDR threshold used to define a significant association with accelerometer-derived MVPA. To assess the ability of guideline-adherent activity to stratify risk of incident disease, we plotted the 5-year cumulative risk of obstructive chronic bronchitis, hypertension, type 2 diabetes, and gastritis/duodenitis (four exemplar conditions having significant associations with both accelerometer-derived and inferred MVPA), stratified at the guideline-based threshold of ≥150 minutes/week of MVPA. We also generated adjusted risk curves by plotting the predicted risk outputs from stratified Cox models fitted with the average age of the sample, guideline-adherent activity as a stratification variable, and male versus female sex separately.

We performed several sensitivity analyses to assess the robustness of our findings. First, we fit analogous models assessing for associations between measured and genetically inferred MVPA and incident disease within age subgroups (i.e., <55, 55-64, and ≥65 years), approximating tertiles of the sample distribution. Second, we assessed associations between measured and inferred MVPA meeting alternative cutoffs of activity (i.e., ≥75 minutes and ≥300 minutes of MVPA/week), with the latter threshold representing activity levels recommended by the World Health Organization for additional health benefit.^4^ Third, to assess whether associations between inferred activity and incident disease may have been affected by reverse causation mediated by horizontal pleiotropy in the genetic instrument (i.e., disease associations arising from variants associated with existing disease which are also associated with lower activity), we re-tested associations with selected exemplar diseases having strong associations in the primary analyses using a genetic instrument developed among individuals without any of the conditions at the time of blood sample collection.

Except where otherwise specified, all analyses were performed using R v4.0^41^ (packages: ‘survival’, ‘data.table’, ‘fdrtool’). Genome-wide association results were evaluated at p=5×10^−8^. Otherwise, all p-values thresholds were corrected for multiplicity by targeting a false discovery rate of 1%. P-values rounding to zero at maximum precision are displayed as 1×10^−10^.

## Data Availability

UK Biobank data are freely available for research purposes by application (https://www.ukbiobank.ac.uk/). Data processing scripts used to perform the analyses described herein can be found at (https://github.com/shaankhurshid/acceleration_phewas). Detailed summary data supporting major study results are provided in the Supplementary Data.

