## Supplemental Material for "Accelerometer-Derived and Genetically Inferred Physical Activity and Human Disease"


| Page | Item |
|---|---|
| 1 | **Table 1.** Moderate-to-vigorous physical activity genome-wide significant and suggestive loci |
| 2 | **Table 2.** Polygenic risk score performance for moderate-to-vigorous physical activity |
| 3 | **Table 3.** Associations between genetically inferred activity and incident disease with and without inclusion of prevalent diseases in the discovery genome-wide association study |
| 4 | **Figure 1.** Distribution of measured and genetically inferred MVPA |
| 5 | **Figure 2.** Overview of analysis samples |
| 6 | **Figure 3.** Strongest associations between measured and genetically inferred MVPA and incident disease |
| 8 | **Figure 4.** Associations between accelerometer-derived guideline-adherent physical activity and incident disease |
| 9 | **Figure 5.** Associations between mean acceleration and incident disease |
| 10 | **Figure 6.** Associations between self-reported MVPA and incident disease |
| 11 | **Figure 7.** Associations between self-reported guideline-adherent activity and incident disease |
| 12 | **Figure 8.** Total count of diseases within each category having significant associations with guideline-adherent physical activity |
| 13 | **Figure 9.** Proportion of diseases within each category having significant associations with physical activity |
| 14 | **Figure 10.** Accelerometer-derived moderate-to-vigorous physical activity quantile-quantile plot |
| 15 | **Figure 11.** Accelerometer-derived moderate-to-vigorous activity Manhattan plot |
| 16 | **Figure 12.** Associations between genetically inferred guideline-adherent activity and incident disease |
| 17 | **Figure 13.** Age-adjusted cumulative risk of disease stratified by level of physical activity in men |
| 18 | **Figure 14.** Age-adjusted cumulative risk of disease stratified by level of physical activity in women |

19 **Figure 15.** Associations between measured MVPA and incident disease across subgroups of age

20 **Figure 16.** Associations between genetically inferred MVPA and incident disease across subgroups of age

21 **Figure 17.** Associations between measured MVPA and incident disease using varying thresholds

22 **Figure 18.** Associations between genetically inferred MVPA and incident disease using varying thresholds

23 **Supplemental References**

**Table 1**. Moderate-to-vigorous physical activity genome-wide significant and suggestive loci

| SNP | Chromosome Position | Effect allele | Non-effect allele | Effect allele frequency | Beta | Standard error | p | Nearest gene | Function |
|---|---|---|---|---|---|---|---|---|---|
| *Genome-wide significant ($p < 5x10^{-8}$)* | | | | | | | | | |
| rs7098100 | 10:21834536 | G | A | 0.66 | 0.076 | 0.011 | $1.60x10^{-12}$ | *MLLT10* | Intron |
| rs11165468 | 1:96160850 | T | C | 0.58 | 0.061 | 0.010 | $4.20x10^{-09}$ | *RWDD3* | Intron |
| *Suggestive ($p < 1x10^{-6}$)* | | | | | | | | | |
| rs4324657 | 5:78862560 | C | A | 0.93 | -0.107 | 0.020 | $1.20x10^{-07}$ | *TENT2* | Intron |
| rs17789849 | 2:24571489 | G | A | 0.92 | -0.098 | 0.019 | $2.30x10^{-07}$ | *ITSN2* | Intron |
| rs61991641 | 14:77502798 | C | T | 0.66 | 0.055 | 0.011 | $2.70x10^{-07}$ | *IRF2BPL* | Upstream transcript |
| rs79348667 | 1:51540509 | T | A | 0.99 | -0.226 | 0.045 | $4.50x10^{-07}$ | *C1orf185* | - |
| rs181092050 | 2:38361804 | T | A | 0.96 | -0.130 | 0.026 | $5.30x10^{-07}$ | *CYP1B1* | Intron |
| rs62263330 | 3:86140222 | T | A | 0.62 | -0.054 | 0.011 | $7.10x10^{-07}$ | *CADM2* | - |
| rs72755809 | 5:25035599 | C | T | 0.72 | -0.057 | 0.011 | $7.40x10^{-07}$ | *CDH10* | - |
| rs7402884 | 15:77381959 | A | G | 0.13 | 0.076 | 0.015 | $7.60x10^{-07}$ | *TSPAN3* | - |
| rs390316 | 14:78596887 | A | C | 0.38 | -0.052 | 0.011 | $7.80x10^{-07}$ | *NRXN3* | Upstream transcript |
| - | 7:98255668 | CTGTT | C | 0.95 | -0.115 | 0.023 | $8.50x10^{-07}$ | *NPTX2* | - |

**Table 2**. Polygenic risk score performance for moderate-to-vigorous physical activity

| Score | P1* | P2* | $R^{2}$* | # variants | $R^2$ for MVPA† |
|---|---|---|---|---|---|
| 1 | 0.0001 | 0.01 | 0.5 | 612 | 0.064 |
| 2 | 0.000001 | 0.01 | 0.5 | 13 | 0.0029 |
| 3 | 0.000001 | 0.01 | 0.3 | 11 | 0.0030 |
| 4 | 0.0001 | 0.01 | 0.3 | 561 | 0.066 |

*P1 = primary p-value threshold used to select variants, P2 = secondary p-value threshold used to select variants, $R^2$ = information threshold (variants sufficiently informative for selected variants are considered candidate proxies for clumping procedure)
†Reflects the $R^2$ from a univariable linear regression of square root-transformed MVPA on each respective square root-transformed MVPA PRS

**Table 3.** Associations between genetically inferred activity and incident disease with and without inclusion of prevalent diseases in the discovery genome-wide association study

| | | *Primary analysis* | | *Excluding prevalent diseases** | |
|---|---|---|---|---|---|
| **Disease** | **Category** | **Hazard ratio per 1-SD increase (95% CI)** | **p** | **Hazard ratio per 1-SD increase (95% CI)** | **p** |
| Obstructive chronic bronchitis | Respiratory | 0.82 (0.79-0.85) | $1.0 \times 10^{-10}$ | 0.84 (0.82-0.87) | $3.3 \times 10^{-16}$ |
| Peripheral vascular disease | Circulatory | 0.92 (0.88-0.95) | $5.1 \times 10^{-6}$ | 0.94 (0.91-0.97) | $6.7 \times 10^{-4}$ |
| Type 2 diabetes | Endocrine/metabolic | 0.92 (0.90-0.94) | $1.0 \times 10^{-10}$ | 0.95 (0.93-0.97) | $8.4 \times 10^{-8}$ |
| Gastritis/duodenitis | Digestive | 0.94 (0.92-0.96) | $3.8 \times 10^{-13}$ | 0.97 (0.95-0.98) | $2.9 \times 10^{-5}$ |

*Depicts results obtained using genetically inferred moderate-vigorous physical activity (MVPA) estimated using a genetic instrument developed from a genome-wide association study excluding all individuals diagnosed with any of the four conditions listed at the time of blood sample collection
SD = standard deviation

**Figure 1**. Distribution of measured and genetically inferred MVPA

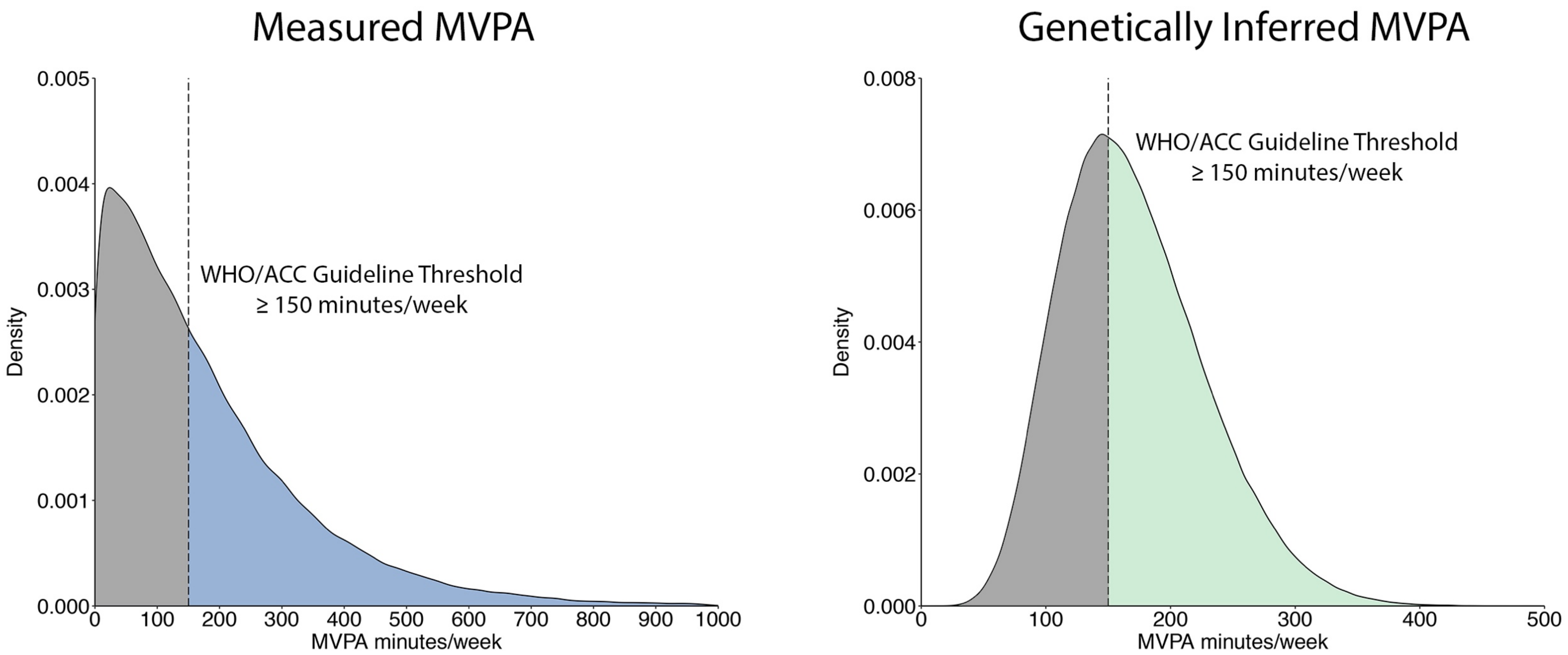


Depicted are the distributions of accelerometer-derived moderate-to-vigorous physical activity (MVPA, left) and genetically inferred MVPA (right). The left plot represents the accelerometer sample (N=96,466) and the right plot represents the Mendelian randomization sample (N=392,058). In both plots, the hashed vertical line represents the level of MVPA recommended by consensus activity guidelines (i.e., ≥150 minutes/week[1–3]), and the grayed area represents individuals whose activity is below the threshold.

**Figure 2**. Overview of analysis samples

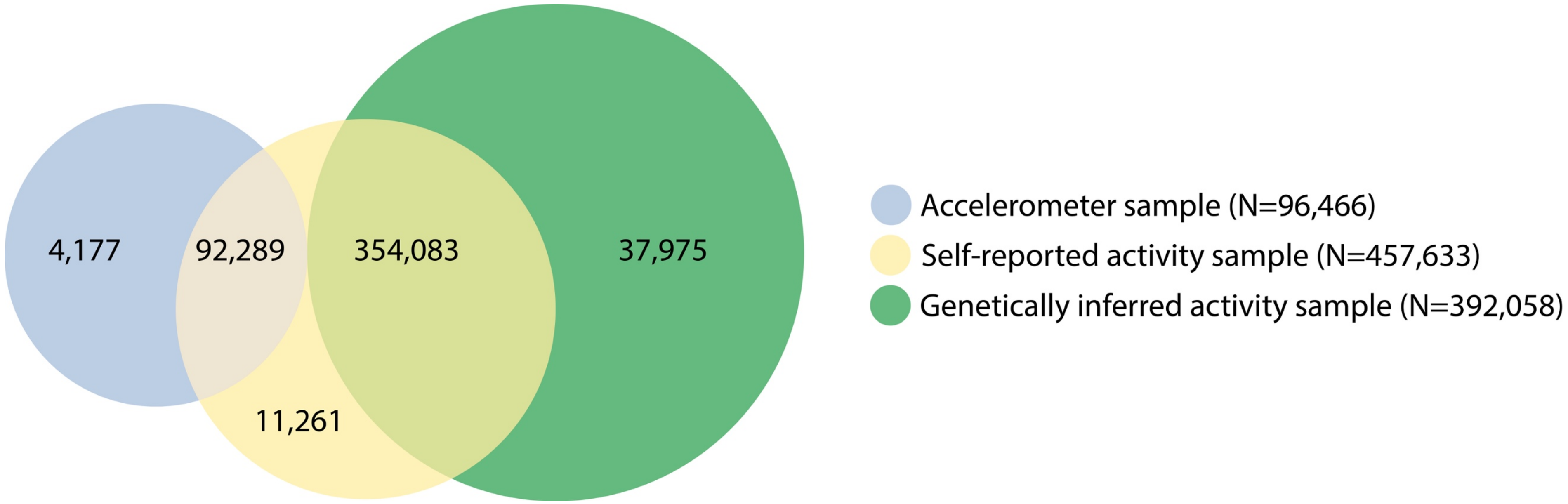


Depicted is a summary of the analysis samples, including counts of overlapping individuals where applicable. A total of 96,466 individuals were included in the accelerometer analysis (blue), with no overlap between those individuals and the genetically inferred activity sample (green). A proportion of both the accelerometer and the genetically inferred activity samples provided self-reported activity for analysis (yellow).

**Figure 3.** Strongest associations between measured and genetically inferred MVPA and incident disease

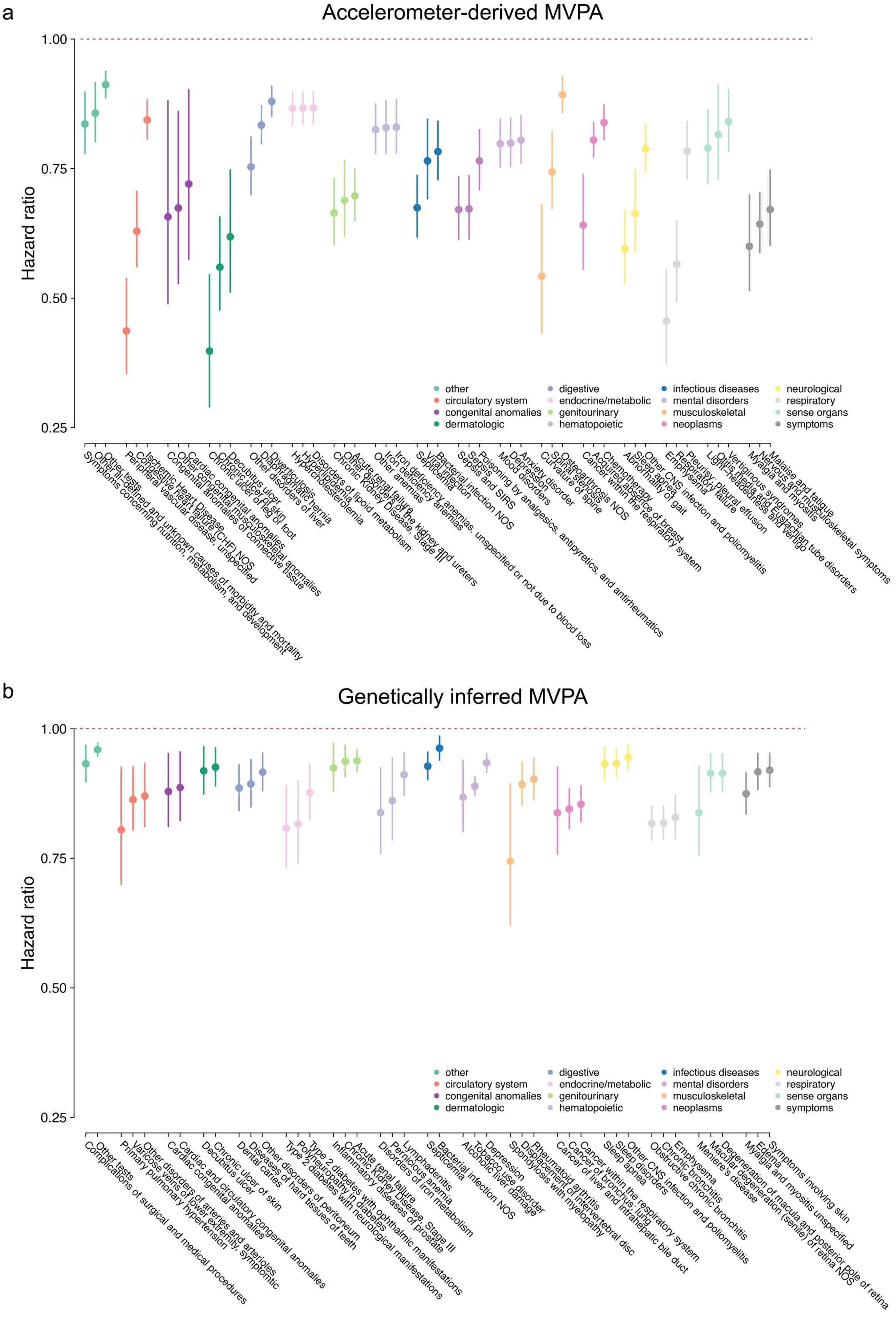

Each plot depicts the top three conditions in each disease category having the strongest associations with accelerometer-derived moderate-vigorous physical activity (MVPA, **panel a**) and genetically inferred MVPA (**panel b**). Specific diseases are identified by name below each plot. Only associations significant at a false discovery rate of 1% are shown. Diseases are colored by category (see legends).

**Figure 4**. Associations between accelerometer-derived guideline-adherent physical activity and incident disease

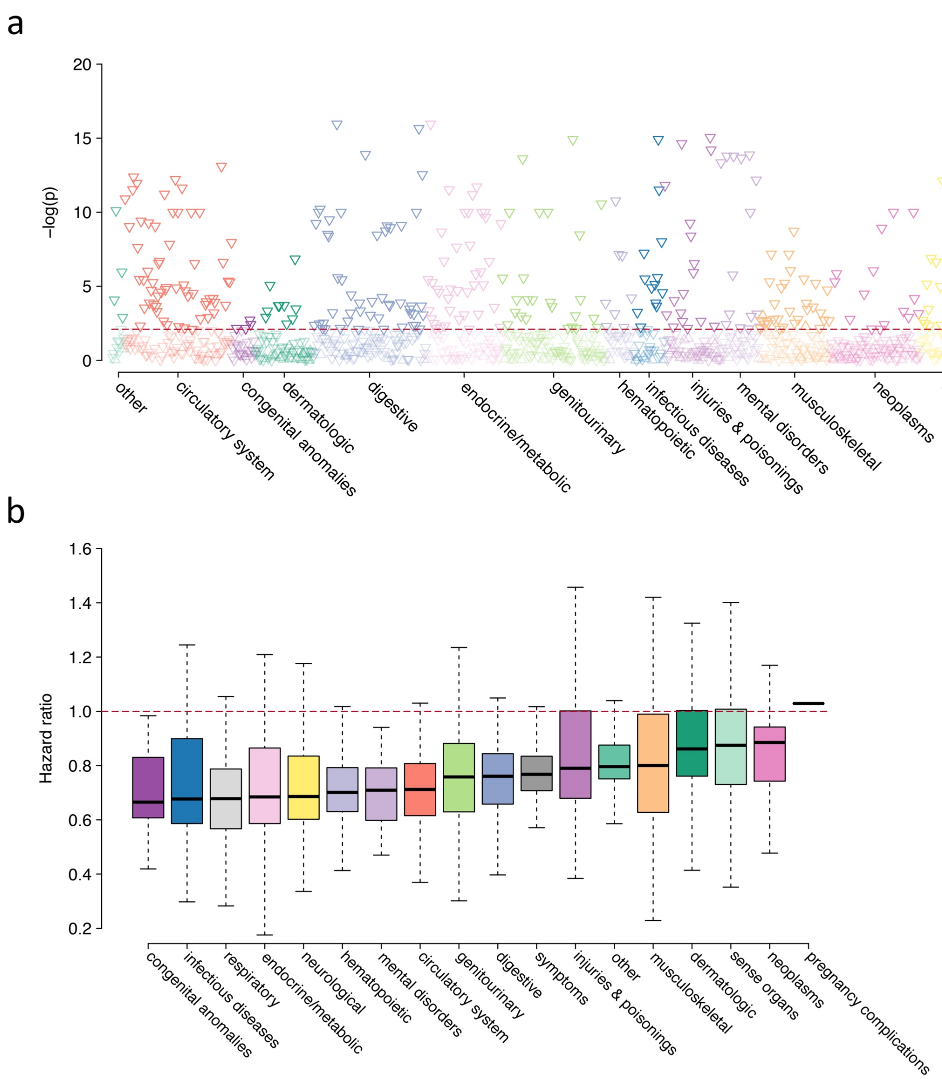


Depicted are the results of phenome-wide disease association testing with accelerometer-derived activity meeting guideline recommendations (≥150 minutes of moderate-vigorous physical activity per week[1–3]) as the exposure of interest, in Cox proportional hazards models adjusted for age, sex, and body mass index. **Panel a** plots the negative log10 p-value for the association between guideline-adherent activity and each individual disease (grouped by category on the x-axis), with darker shaded points meeting significance at a false discovery rate of 1% (threshold depicted by horizontal dashed red line). Upward facing triangles represent increased risk (hazard ratios > 1), while downward facing triangles represent reduced risk (hazard ratio < 1). **Panel b** shows the distribution of hazard ratios observed in the presence of guideline-adherent activity across each disease category (x-axis), with the thick horizontal line depicting the median hazard ratio within the category, the box representing quartile 1 to quartile 3, and the whiskers extending 1.5 interquartile ranges beyond the box. Categories are arranged by increasing median hazard ratio, from lowest (left) to highest (right).

**Figure 5**. Associations between mean acceleration and incident disease

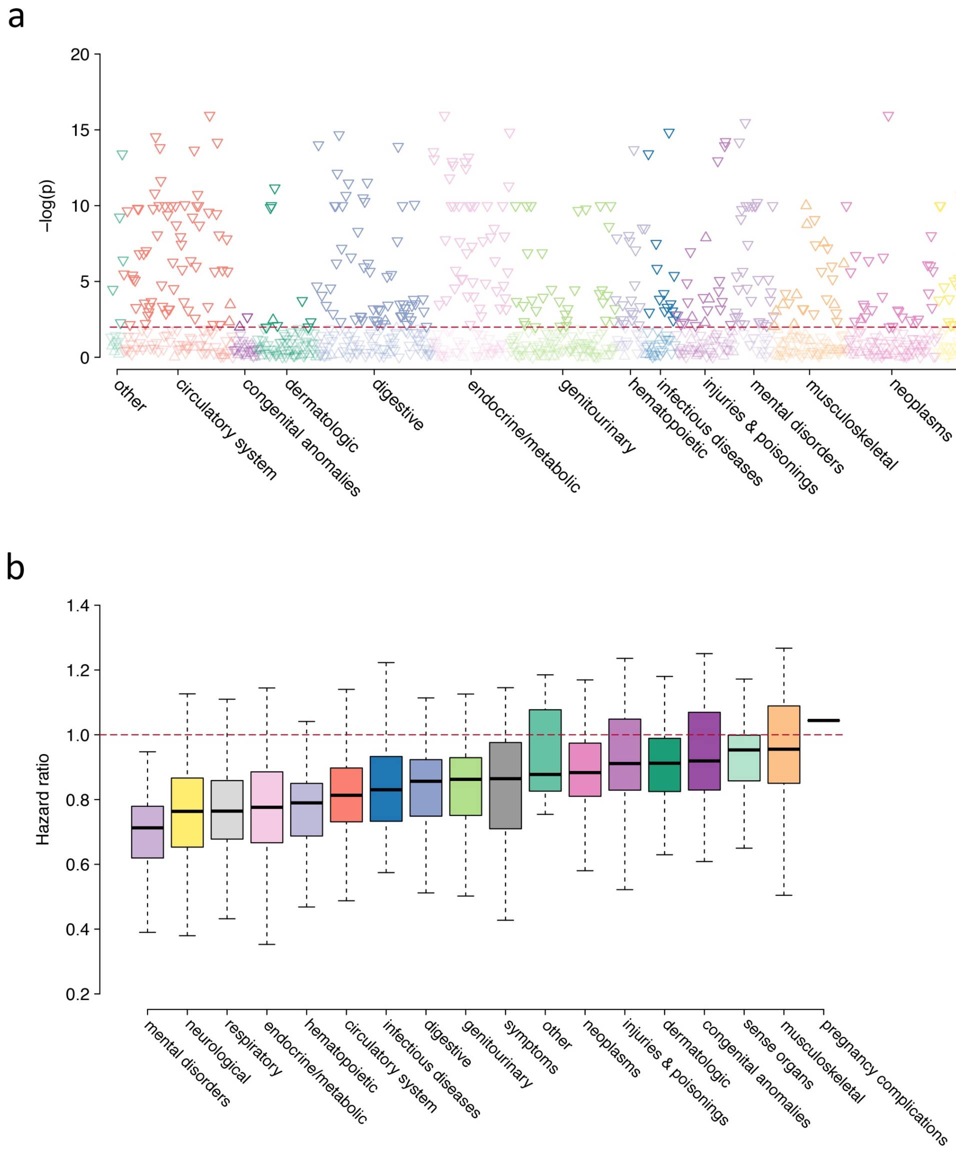


Depicted are the results of phenome-wide disease association testing with mean acceleration as the exposure of interest, in Cox proportional hazards models adjusted for age, sex, and body mass index. **Panel a** plots the negative log10 p-value for the association between mean acceleration and each individual disease (grouped by category on the x-axis), with darker shaded points meeting significance at a false discovery rate of 1% (threshold depicted by horizontal dashed red line). Upward facing triangles represent increased risk (hazard ratios > 1), while downward facing triangles represent reduced risk (hazard ratio < 1). **Panel b** shows the distribution of hazard ratios observed per 1 standard deviation increase in mean acceleration across each disease category (x-axis), with the thick horizontal line depicting the median hazard ratio within the category, the box representing quartile 1 to quartile 3, and the whiskers extending 1.5 interquartile ranges beyond the box. Categories are arranged by increasing median hazard ratio, from lowest (left) to highest (right).

**Figure 6**. Associations between self-reported MVPA and incident disease

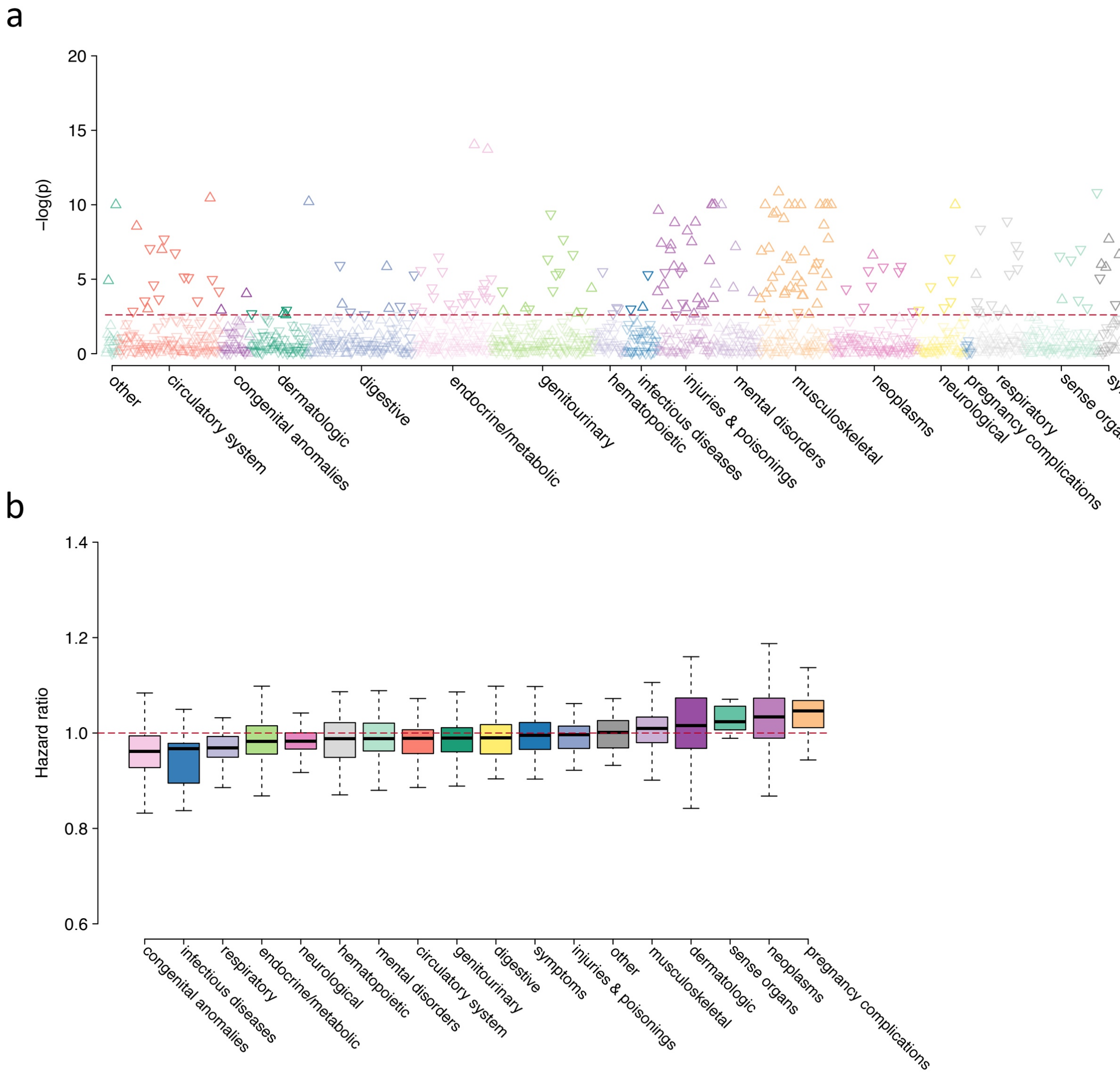


Depicted are the results of phenome-wide disease association testing with self-reported moderate-to-vigorous physical activity (MVPA) as the exposure of interest, in Cox proportional hazards models adjusted for age, sex, and body mass index. **Panel a** plots the negative log10 p-value for the association between self-reported MVPA and each individual disease (grouped by category on the x-axis), with darker shaded points meeting significance at a false discovery rate of 1% (threshold depicted by horizontal dashed red line). Upward facing triangles represent increased risk (hazard ratios > 1), while downward facing triangles represent reduced risk (hazard ratio < 1). **Panel b** shows the distribution of hazard ratios observed for every 1 standard deviation increase in self-reported MVPA across each disease category (x-axis), with the thick horizontal line depicting the median hazard ratio within the category, the box representing quartile 1 to quartile 3, and the whiskers extending 1.5 interquartile ranges beyond the box. Categories are arranged by increasing median hazard ratio, from lowest (left) to highest (right).

**Figure 7**. Associations between self-reported guideline-adherent activity and incident disease

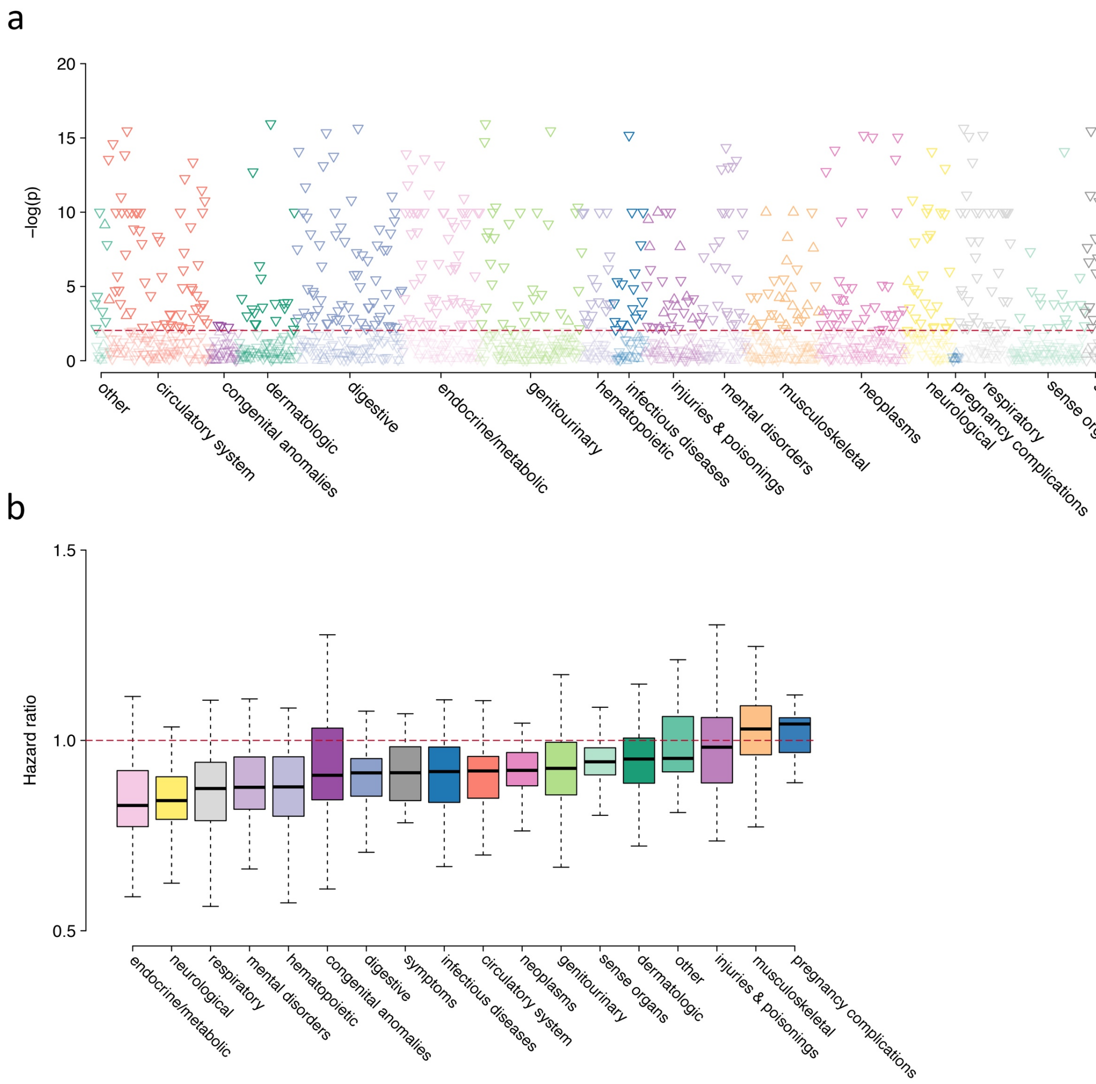


Depicted are the results of phenome-wide disease association testing with self-reported activity meeting guideline recommendations (≥150 minutes of moderate-vigorous physical activity per week[1–3]) as the exposure of interest, in Cox proportional hazards models adjusted for age, sex, and body mass index. **Panel a** plots the negative log10 p-value for the association between self-reported guideline-adherent activity and each individual disease (grouped by category on the x-axis), with darker shaded points meeting significance at a false discovery rate of 1% (threshold depicted by horizontal dashed red line). Upward facing triangles represent increased risk (hazard ratios > 1), while downward facing triangles represent reduced risk (hazard ratio < 1). **Panel b** shows the distribution of hazard ratios observed in the presence of guideline-adherent activity across each disease category (x-axis), with the thick horizontal line depicting the median hazard ratio within the category, the box representing quartile 1 to quartile 3, and the whiskers extending 1.5 interquartile ranges beyond the box. Categories are arranged by increasing median hazard ratio, from lowest (left) to highest (right).

**Figure 8.** Total count of diseases within each category having significant associations with guideline-adherent physical activity

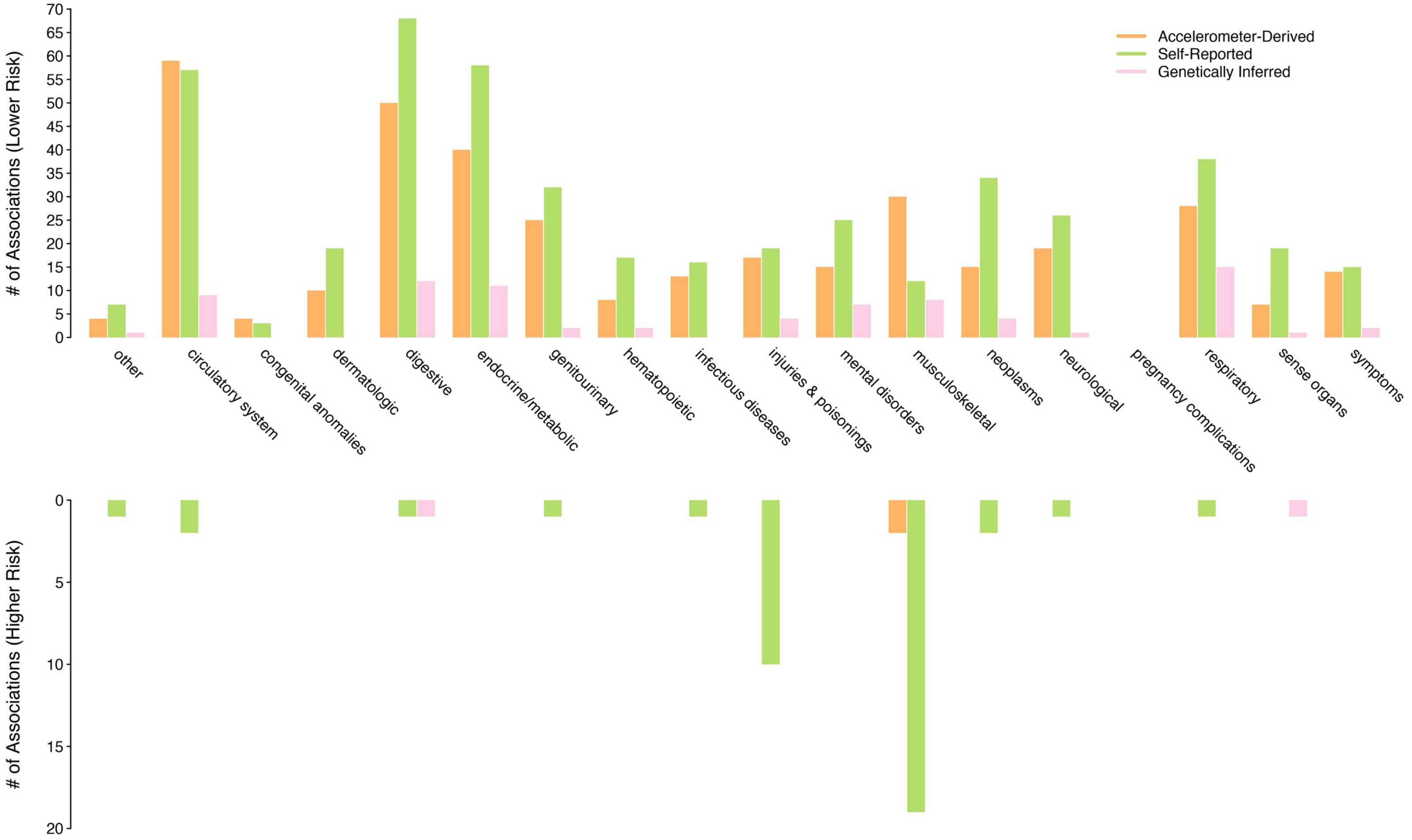


Depicted is the total count of diseases within each category having a significant association with guideline-adherent physical activity (i.e., ≥150 minutes of moderate-vigorous physical activity per week[1–3]). Three sources for classifying activity are compared (see legend). The top panel depicts associations indicating reduced disease risk (i.e., hazard ratios less than one), while the bottom panel depicts associations indicating increased disease risk (i.e., hazard ratios greater than one). Only associations significant at a false discovery rate of 1% are depicted.

**Figure 9.** Proportion of diseases within each category having significant associations with physical activity

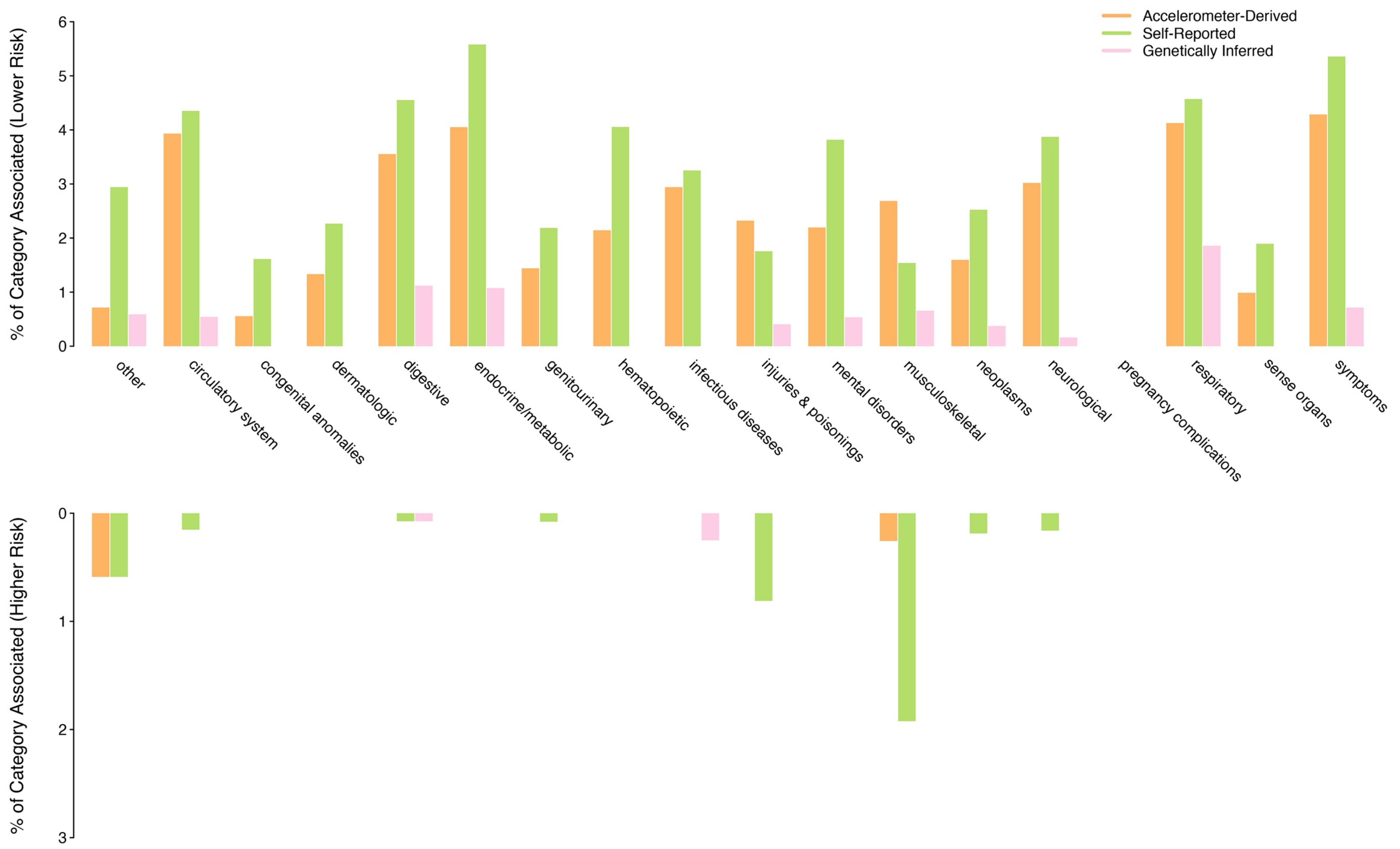


Depicted is the proportion of diseases within each category having a significant association with physical activity. Five different physical activity classification methods are compared (see legend). The top panel depicts associations indicating reduced disease risk (i.e., hazard ratios less than one), while the bottom panel depicts associations indicating increased disease risk (i.e., hazard ratios greater than one). Only diseases meeting phenome-wide significance at a false discovery rate of 1% are depicted.

**Figure 10.** Accelerometer-derived moderate-to-vigorous physical activity quantile-quantile plot

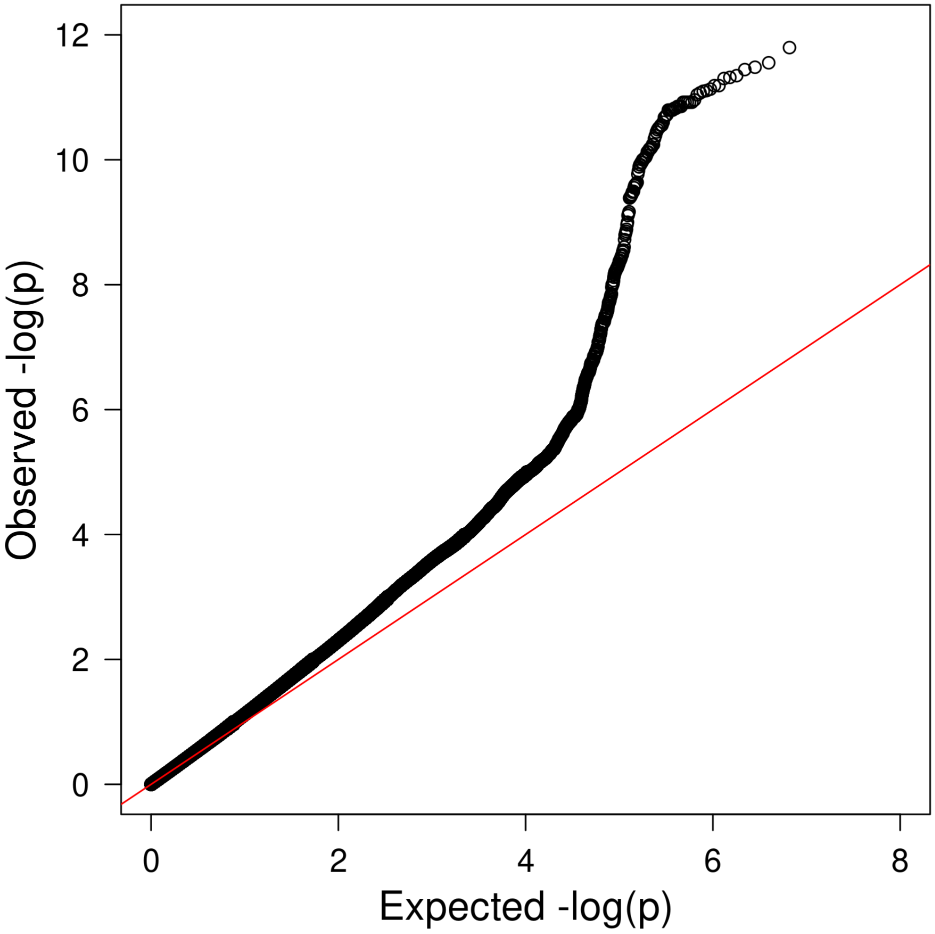


Depicted is a quantile-quantile plot of a genome-wide association study of square root-transformed accelerometer-derived moderate-vigorous physical activity. The genomic control factor was 1.15 with a linkage-disequilibrium score regression intercept of 1.008, consistent with polygenicity rather than inflation.[4]

**Figure 11**. Accelerometer-derived moderate-to-vigorous activity Manhattan plot

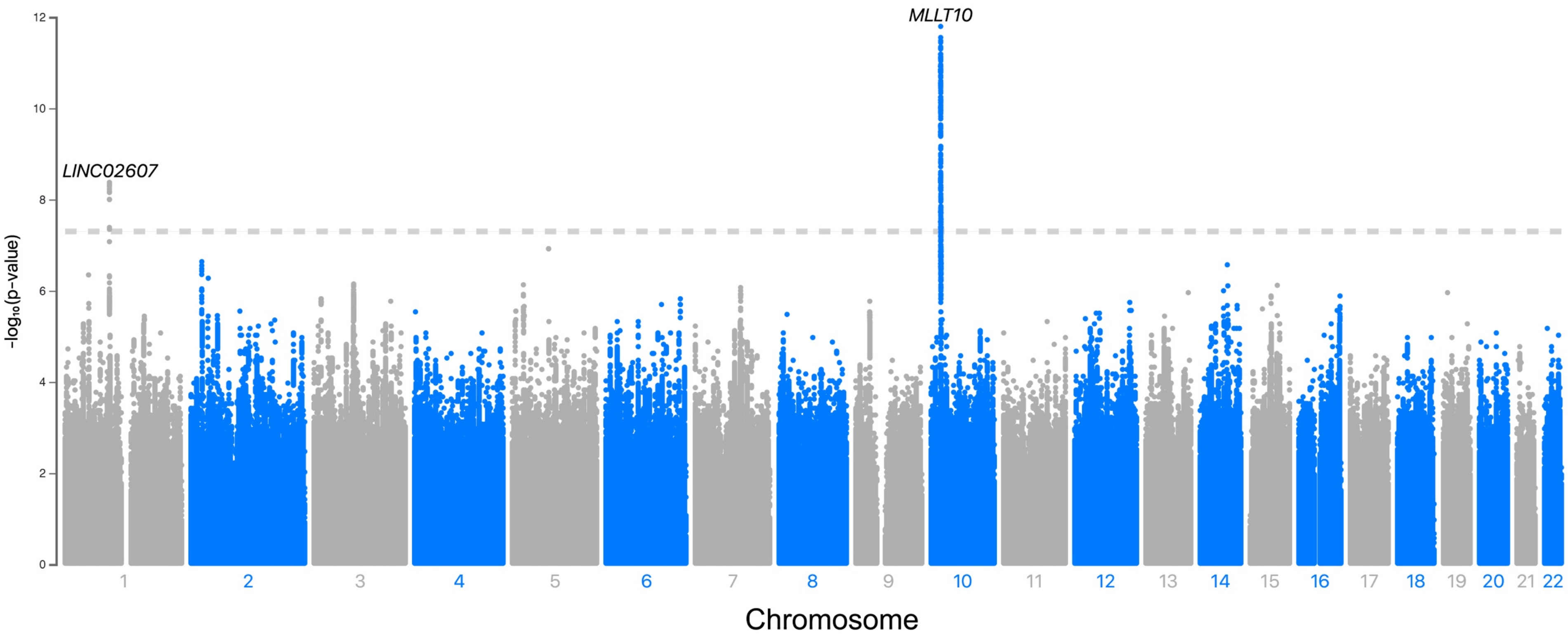


Depicted are the results of a genome-wide association study of accelerometer-derived moderate-vigorous physical activity. A square-root transform was applied to attenuate right skew in the distribution (see main text). Only variants having minor allele frequency ≥1% are depicted. The hashed gray line depicts the standard genome-wide significance level ($p=5x10^{-8}$).

**Figure 12.** Associations between genetically inferred guideline-adherent activity and incident disease

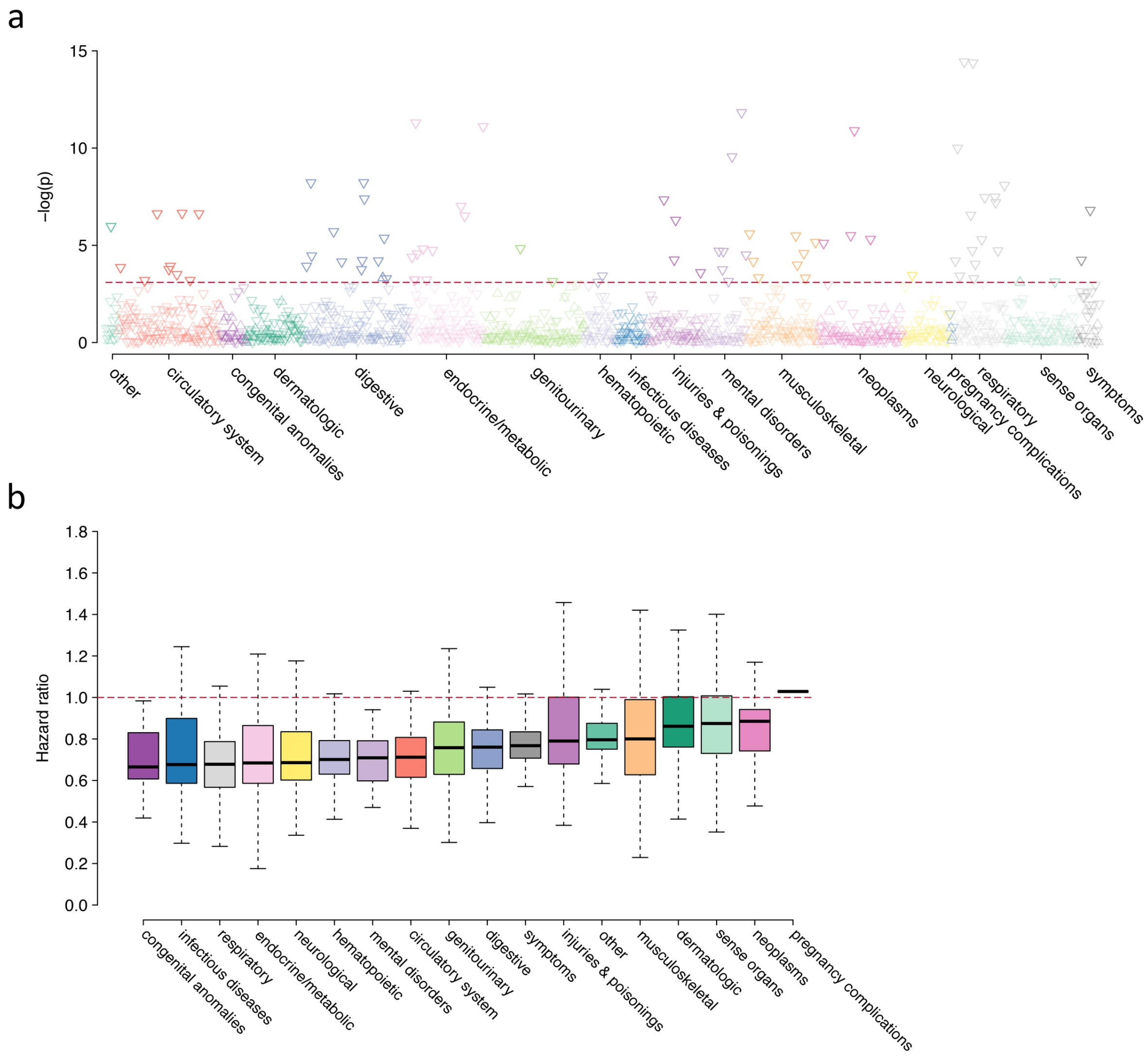


Depicted are the results of phenome-wide disease association testing with genetically inferred activity meeting guideline recommendations (≥150 minutes of moderate-vigorous physical activity per week[1–3]) as the exposure of interest, in Cox proportional hazards models adjusted for age and sex. **Panel a** plots the negative log10 p-value for the association between guideline-adherent activity and each individual disease (grouped by category on the x-axis), with darker shaded points meeting significance at a false discovery rate of 1% (threshold depicted by horizontal dashed red line). Upward facing triangles represent increased risk (hazard ratios > 1), while downward facing triangles represent reduced risk (hazard ratio < 1). **Panel b** shows the distribution of hazard ratios observed in the presence of guideline-adherent activity across each disease category (x-axis), with the thick horizontal line depicting the median hazard ratio within the category, the box representing quartile 1 to quartile 3, and the whiskers extending 1.5 interquartile ranges beyond the box. Categories are arranged by increasing median hazard ratio, from lowest (left) to highest (right).

**Figure 13.** Age-adjusted cumulative risk of disease stratified by level of physical activity in men

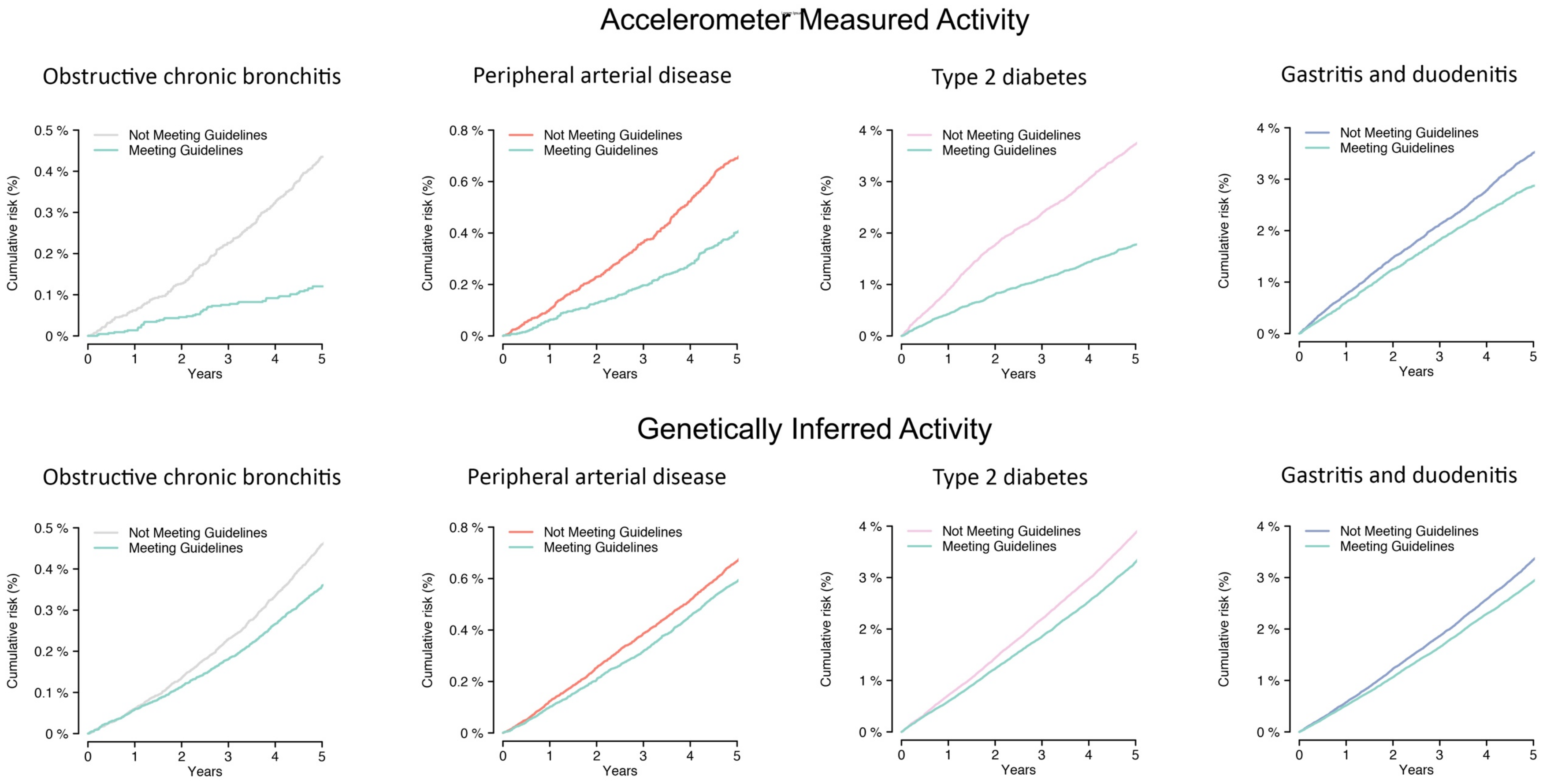


Depicted is the age-adjusted 5-year cumulative risk of obstructive chronic bronchitis, peripheral arterial disease, type 2 diabetes, and gastritis/duodenitis for men, stratified by guideline-adherent activity (≥150 minutes of MVPA/week[4–6]) according to accelerometer-derived MVPA (top panels) and according to genetically inferred MVPA (bottom). Curves are derived from stratified Cox models with each respective disease as the outcome, age and sex as covariates, and guideline-adherent activity as a stratification variable. Representative diseases were selected from the four categories having the greatest enrichment for associations with activity, where each disease was significantly associated with both accelerometer-derived and genetically inferred activity at a false discovery rate of 1%.

**Figure 14.** Age-adjusted cumulative risk of disease stratified by level of physical activity in women

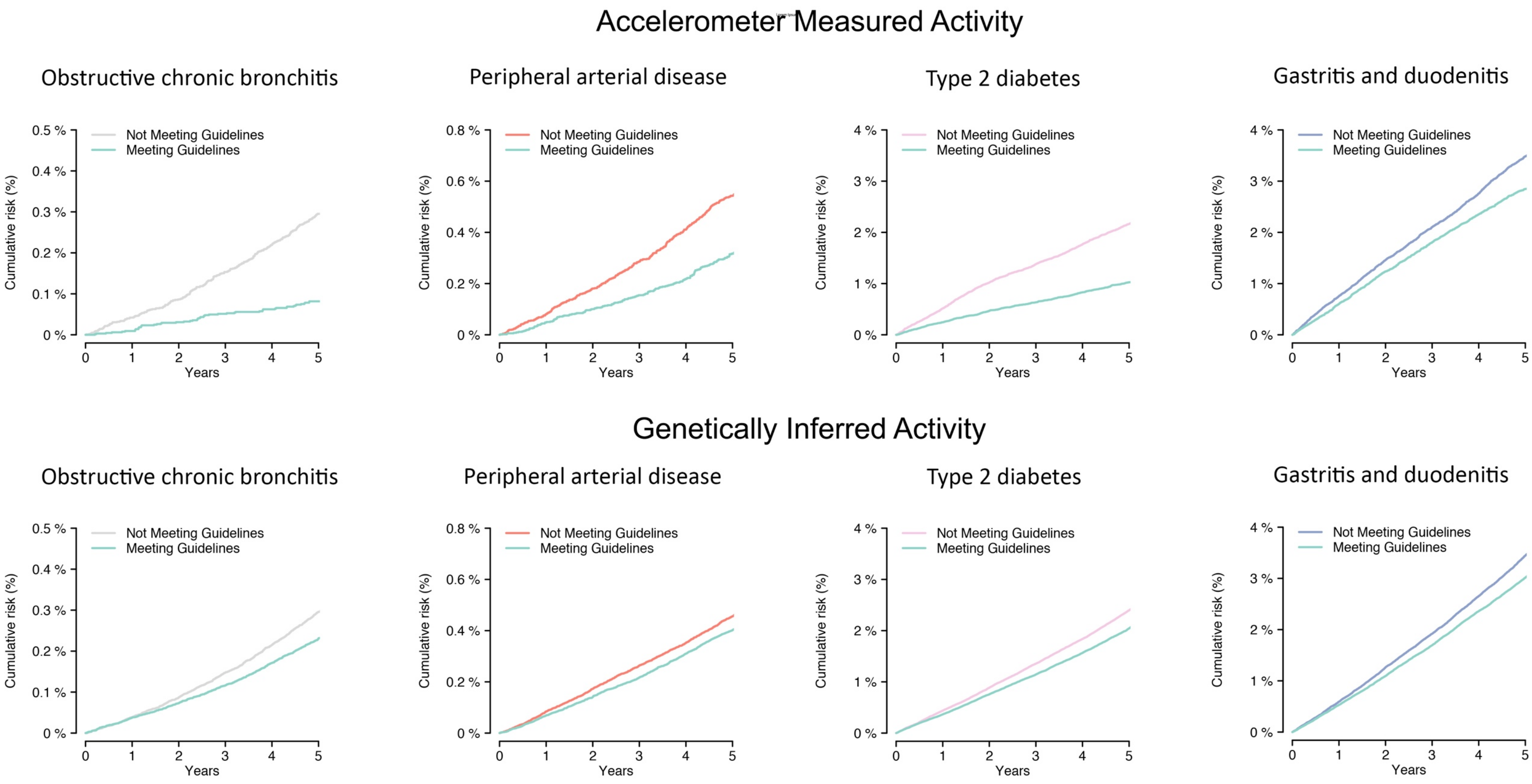


Depicted is the age-adjusted 5-year cumulative risk of obstructive chronic bronchitis, peripheral arterial disease, type 2 diabetes, and gastritis/duodenitis for women, stratified by guideline-adherent activity (≥150 minutes of MVPA/week[4–6]) according to accelerometer-derived MVPA (top panels) and according to genetically inferred MVPA (bottom). Curves are derived from stratified Cox models with each respective disease as the outcome, age and sex as covariates, and guideline-adherent activity as a stratification variable. Representative diseases were selected from the four categories having the greatest enrichment for associations with activity, where each disease was significantly associated with both accelerometer-derived and genetically inferred activity at a false discovery rate of 1%.

**Figure 15.** Associations between measured MVPA and incident disease across subgroups of age

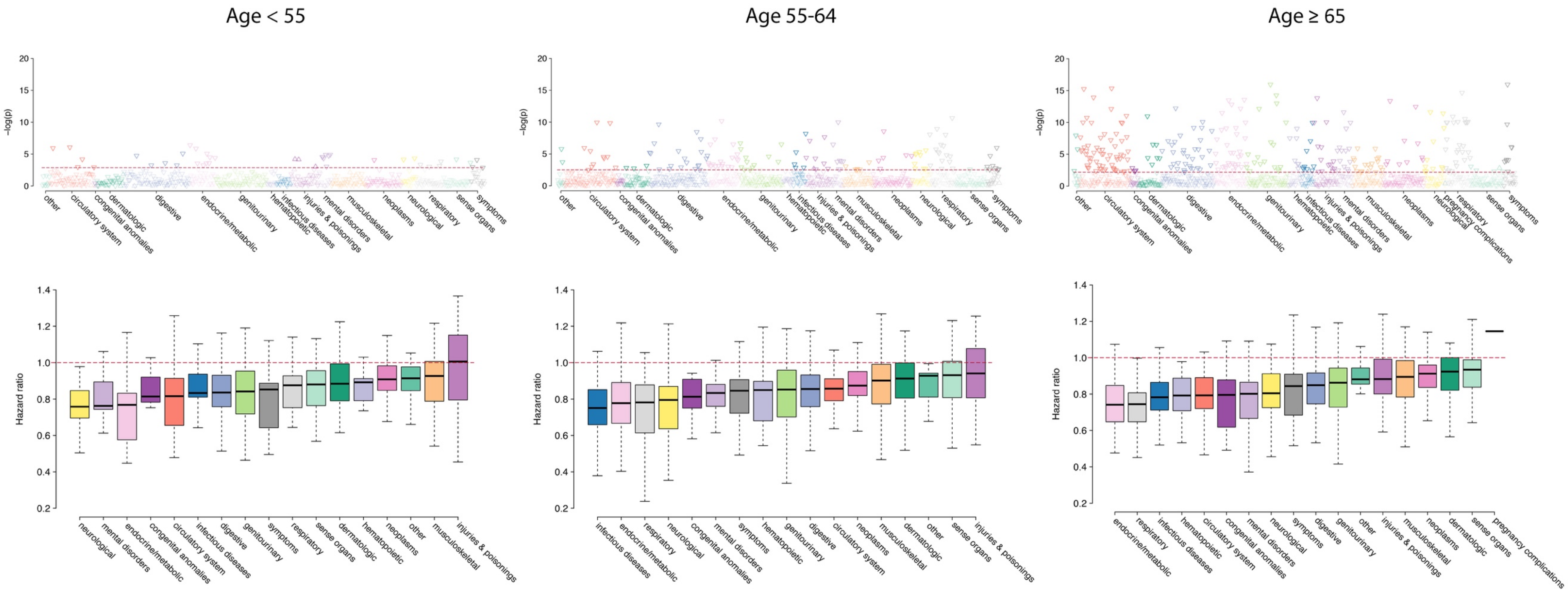


Depicted are associations between accelerometer-derived moderate-vigorous physical activity (MVPA) and incident disease across subgroups of age. Age cutoffs were chosen to approximate tertiles of the sample distribution (subgroup n for age <55: 20,568, age 55-65: 33,878, age ≥ 65: 42,020). Upper panels depict the negative log10 p-value for the association between MVPA and each individual disease (grouped by category on the x-axis), with darker shaded points meeting significance at a false discovery rate of 1% (threshold depicted by horizontal dashed red line). Upward facing triangles represent increased risk (hazard ratios > 1), while downward facing triangles represent reduced risk (hazard ratio < 1). Lower panels depict the distribution of hazard ratios observed per 1-standard deviation increase in MVPA across each disease category (x-axis), with the thick horizontal line depicting the median hazard ratio within the category, the box representing quartile 1 to quartile 3, and the whiskers extending 1.5 interquartile ranges beyond the box. Categories are arranged by increasing median hazard ratio, from lowest (left) to highest (right).

**Figure 16.** Associations between genetically inferred MVPA and incident disease across subgroups of age

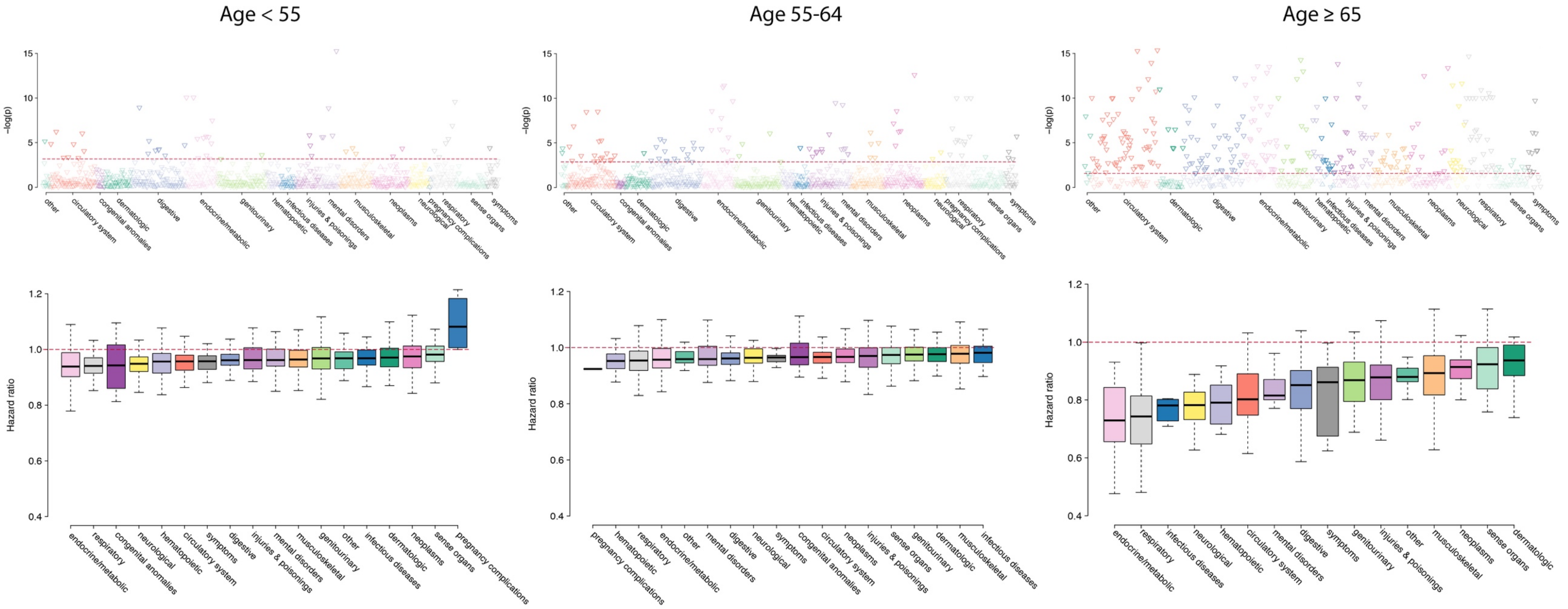


Depicted are associations between genetically inferred moderate-vigorous physical activity (MVPA) and incident disease across subgroups of age. Age cutoffs were chosen to approximate tertiles of the sample distribution (subgroup n for age <55: 150,299, age 55-65: 163,471, age ≥ 65: 78,288). Upper panels depict the negative log10 p-value for the association between MVPA and each individual disease (grouped by category on the x-axis), with darker shaded points meeting significance at a false discovery rate of 1% (threshold depicted by horizontal dashed red line). Upward facing triangles represent increased risk (hazard ratios > 1), while downward facing triangles represent reduced risk (hazard ratio < 1). Lower panels depict the distribution of hazard ratios observed per 1-standard deviation increase in MVPA across each disease category (x-axis), with the thick horizontal line depicting the median hazard ratio within the category, the box representing quartile 1 to quartile 3, and the whiskers extending 1.5 interquartile ranges beyond the box. Categories are arranged by increasing median hazard ratio, from lowest (left) to highest (right).

**Figure 17.** Associations between measured MVPA and incident disease using varying thresholds

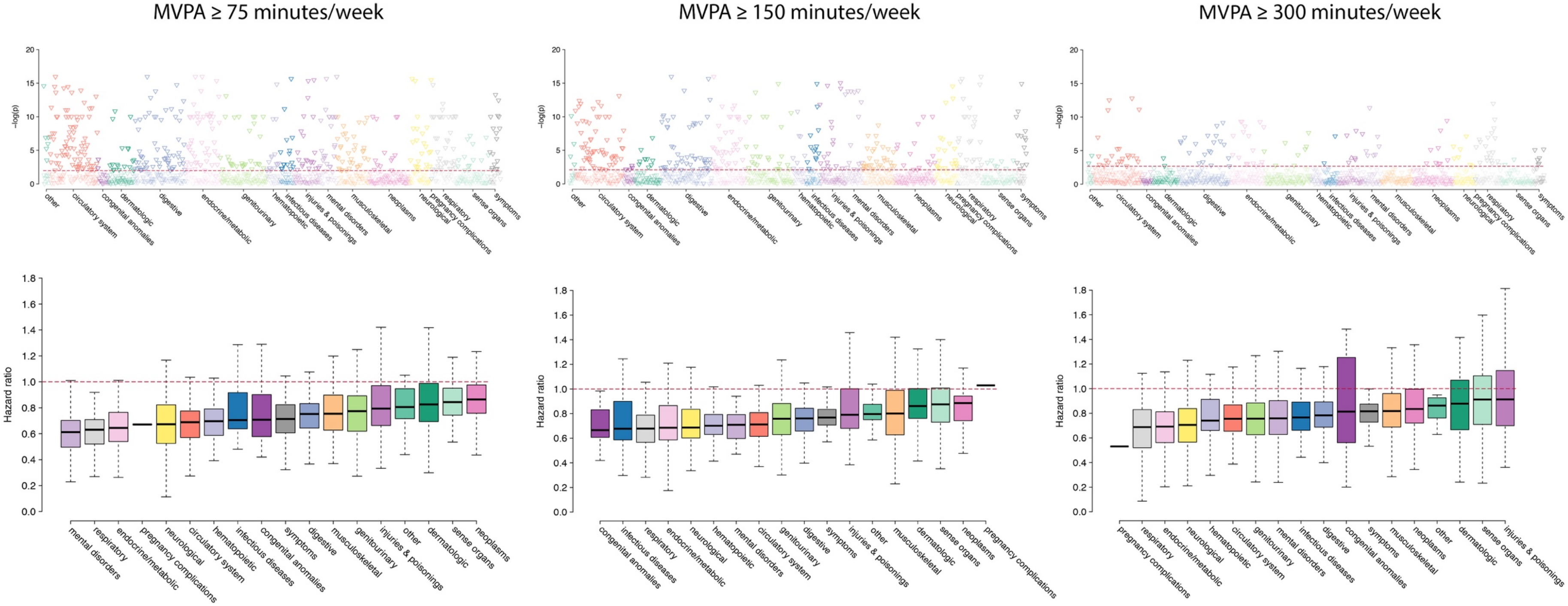


Depicted are associations between measured moderate-vigorous physical activity (MVPA) and incident disease applying varying thresholds. Middle panels depict the standard guideline-based threshold used in the primary analysis (≥150 minutes MVPA/week). Right panels depict the World Health Organization threshold for extended health benefit (≥300 minutes MVPA/week). The left panels depict a lower threshold (≥75 minutes MVPA/week). The number of individuals meeting each threshold are as follows: ≥75 minutes MVPA/week: 67,151 (69.6%), ≥150 minutes MVPA/week: 44,691 (46.3%), ≥300 minutes MVPA/week: 17,873 (18.5%). Upper panels depict the negative log10 p-value for the association between activity meeting the given threshold and each individual disease (grouped by category on the x-axis), with darker shaded points meeting significance at a false discovery rate of 1% (threshold depicted by horizontal dashed red line). Upward facing triangles represent increased risk (hazard ratios > 1), while downward facing triangles represent reduced risk (hazard ratio < 1). Lower panels depict the distribution of hazard ratios observed across each disease category (x-axis), with the thick horizontal line depicting the median hazard ratio within the category, the box representing quartile 1 to quartile 3, and the whiskers extending 1.5 interquartile ranges beyond the box. Categories are arranged by increasing median hazard ratio, from lowest (left) to highest (right).

**Figure 18.** Associations between genetically inferred MVPA and incident disease using varying thresholds

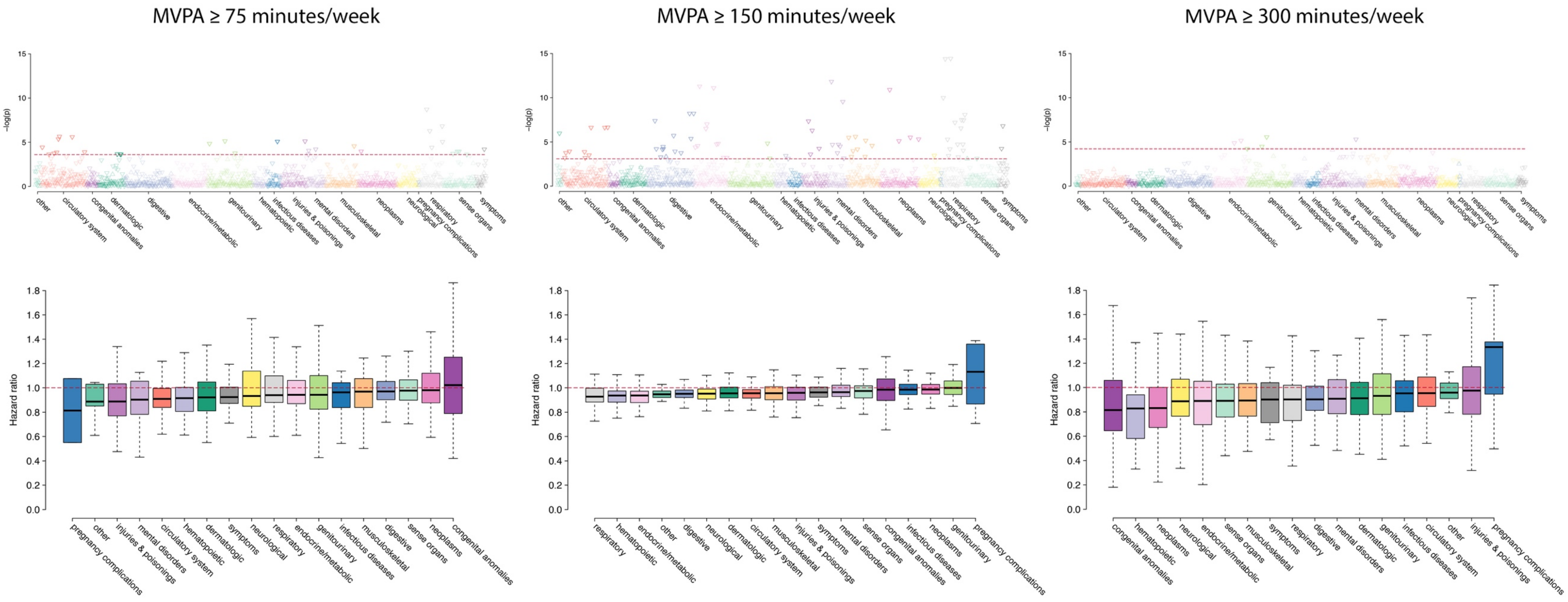


Depicted are associations between measured moderate-vigorous physical activity (MVPA) and incident disease applying varying thresholds. Middle panels depict the standard guideline-based threshold used in the primary analysis (≥150 minutes MVPA/week). Right panels depict the World Health Organization threshold for extended health benefit (≥300 minutes MVPA/week). The left panels depict a lower threshold (≥75 minutes MVPA/week). The number of individuals meeting each threshold are as follows: ≥75 minutes MVPA/week: 383,150 (97.7%), ≥150 minutes MVPA/week: 233,950 (59.7%), ≥300 minutes MVPA/week: 9,221 (2.4%). Upper panels depict the negative log10 p-value for the association between activity meeting the given threshold and each individual disease (grouped by category on the x-axis), with darker shaded points meeting significance at a false discovery rate of 1% (threshold depicted by horizontal dashed red line). Upward facing triangles represent increased risk (hazard ratios > 1), while downward facing triangles represent reduced risk (hazard ratio < 1). Lower panels depict the distribution of hazard ratios observed across each disease category (x-axis), with the thick horizontal line depicting the median hazard ratio within the category, the box representing quartile 1 to quartile 3, and the whiskers extending 1.5 interquartile ranges beyond the box. Categories are arranged by increasing median hazard ratio, from lowest (left) to highest (right).
